# The effect of competition between health opinions on epidemic dynamics

**DOI:** 10.1101/2022.02.10.22270768

**Authors:** Alexandra Teslya, Hendrik Nunner, Vincent Buskens, Mirjam E Kretzschmar

**Author notes:** MEK and VB conceived the study and developed the model. MEK and AT performed stability and bifurcation analysis of the model dynamics. AT implemented the model, carried out all numerical model analyses. AT prepared figures with input from VB, HN, and MEK. All authors participated in the discussion and interpretation of the outputs of the model. AT performed relevant literature overview. AT and MEK wrote the manuscript. All authors contributed to editing of the final version of the manuscript, and gave the final approval for publication. The authors declare no competing interests.

## Abstract

Past major epidemic events showed that when an infectious disease is perceived to cause severe health outcomes, individuals modify health behavior affecting epidemic dynamics. To investigate the effect of this feedback relationship on epidemic dynamics, we developed a compartmental model that couples a disease spread framework with competition of two mutually exclusive health opinions (health-positive and health-neutral) associated with different health behaviors. The model is based on the assumption that individuals switch health opinions as a result of exposure to opinions of others through interpersonal communications. To model opinion switch rates, we considered a family of functions and identified the ones that allow health opinions to co-exist. In the disease-free population, either the opinions cannot co-exist and one of them is always dominating (monobelief equilibrium) or there is at least one stable co-existence of opinions equilibrium. In the latter case, there is multistability between the co-existence equilibrium and the two monobelief equilibria. When two opinions co-exist, it depends on their distribution whether the infection can invade. If presence of the infection leads to increased switching to a health-positive opinion, the epidemic burden becomes smaller than indicated by the basic reproduction number. Additionally, a feedback between epidemic dynamics and health opinion dynamics may result in (sustained) oscillatory dynamics and a switch to a different stable opinion distribution. Our model captures feedback between spread of awareness through social networks and infection dynamics and can serve as a basis for more elaborate individual-based models.

**Significance Statement:** Disease epidemics often co-evolve with opinions on health-related behavior. Most existing models have difficulties understanding co-existence of different opinions in a population when the disease is not present, while we do observe this. We modeled opinion switching process by using an innovative way to capture the dependence of opinion switching rate on the population state. We combined this with network interaction patterns and were able to derive conditions under which a stable co-existence of opinions can occur. We used this insight to explain appearance of epidemic cycles and the population switching between different distributions of opinions. Our work demonstrates that for information interventions accurate understanding of opinion propagation processes is crucial.

---

The notion that the relationship between epidemic dynamics and reactive collective behavior plays an important role in the course of an outbreak of an infectious disease has been recognized in theoretical epidemiology (1–5). This notion is supported by data collected during various outbreaks of infectious diseases, dating back as far as the Spanish flu pandemic of 1918 (1, 3, 5) to SARS pandemic (6, 7) and swine flu pandemic (8), and ending with the ongoing SARS-CoV2 pandemic (9). Two types of societal reactions to an infectious disease outbreak can be distinguished, namely centralized top-down and individual-based bottom-up reactions. First, governing authorities may impose public health interventions aiming at protecting the most vulnerable groups, and mitigating the spread of infection. Typical measures are school closures, limitation of the number of persons in indoor spaces, and travel restrictions. Second, individuals may change their behavior by self-imposing protective measures such as hygiene measures or mask wearing in an effort to defend themselves from infection and its consequences (10). It has been observed that practicing of self-protective measures increased during outbreaks of infectious diseases and declined when the disease was eliminated (6–8). Thus, there is an indication for a feedback relationship between epidemic dynamics and uptake of self-protective measures.

It was not until the 2000s that the importance of this type of reaction for epidemic dynamics was recognized and investigated using mathematical modeling (2, 4, 11). Accounting for the behavior-infection feedback relationships in epidemic models has helped to explain patterns observed in real world data. Multiple epidemic peaks and relatively small outbreaks, where much larger ones were expected, were convincingly shown to be the result of changes in individual human behavior during an epidemic (4, 5).

Health behaviors are a subject to (health) opinion held. The dynamics of circulation of ideas and beliefs in a population is studied in the field known as sociophyics. Even the simplest sociophysics models can have rich dynamics where a number of distinct opinion distributions is possible with a potential for bistability between them (12–14). To understand the effect of the feedback loop between disease spread and health opinion circulation on epidemic dynamics, it is important to understand the role of assumptions about the propagation of opinions on their distribution in the population. In this work we consider the effect of interpersonal communications on the dynamics of health opinion competition using different functional representations for opinion switch rates. We show that depending on the shape of the functional response qualitatively different opinion distributions appear, which in turn affects outlook of an epidemic.

In the context of health-related opinions and the associated self-imposed preventive behaviors, pro- and anti-vaccination sentiments garnered a lot of attention (11, 15–17), while other investigations focused on non-pharmaceutical interventions such as mask wearing and social distancing (2, 4, 9, 18). While ideally vaccination is a nearly instantaneous event that protects an individual for a long time, the latter measures only confer protection while they are being practiced. For emerging infectious diseases for which pharmaceutical interventions are not available, as was the case with COVID-19 in 2020, the extent of the outbreak depends on the uptake rate of non-pharmaceutical measures by the population (19).

Health opinions can fall on a spectrum ranging from health-promoting, adaptors of which practice self-protective measures, to health-indifferent, whereupon individuals having such opinions do not modify their behavior with the aim of protecting their health. The health belief model (10) posits that adopting health promoting measures is motivated by several constructs: (i) perceived susceptibility (risk of contracting a specific health problem), (ii) perceived severity (estimation of the consequences of this problem), (iii) perceived barriers (impediments for adopting a relevant health behavior), (iv) perceived benefits (assessment of effectiveness in avoiding the health problem if the health behavior is adopted), and (v) cues to action (events that bring on adoption of a specific behavior). If an individual believes the disease to be a threat, they may modify their health behavior in a number of ways that affect their susceptibility, the probability of encountering an infectious individual, and duration of infection. In contrast to beliefs, which support adoption of health protective behaviors, individuals may also be indifferent to health-related risks. Indifferent individuals may make little to no effort to protect their health or limit the disease spread. For example, during the AH1N1/09 (“swine flu”) outbreak in 2009, people who were uncertain about the disease and felt that the extent and danger of the outbreak were exaggerated were less likely to change their behavior (20)

Individuals may form and change their opinions when being exposed to communications by a.o. health officials, newscasts, social media, and interpersonal interactions. Ideally, communications by health officials provide accurate information about an epidemic outbreak and possible self-protective measures that individuals can adopt. On the other hand, social media and interpersonal communications can be carriers of misinformation and opinions that may downplay or exaggerate the risks of acquiring infection. Individuals may feel a pressure to conform to their social environment and may adopt an opinion even if it contradicts available evidence or information distributed by health authorities (13). Moreover, by means of digital social media interpersonal communications can spread more widely and rapidly than through the physical contact network, such that the propagation may be stimulated by ongoing communication in media (21).

Here we focus on a health opinion switching process that arises due to interpersonal communication. To investigate the effect of interpersonal communication on the competition of health opinions in the population, we developed a deterministic compartmental model that stratifies the population by opinions. To improve the analytic tractability of the mathematical model, we restrict ourselves to the case of two mutually exclusive opinions, namely health-positive and health-neutral. While health opinions in reality can range on a continuous scale between health awareness and indifference (22), our choice can also be justified by the argument that health related behavior is either practiced or not. So, we assume that holding the health-positive opinion invariably leads to adoption of health protective measures in the face of an outbreak (e.g., mask wearing, increased hands washing, keeping a distance of 1.5 meters from others), while individuals holding the health-neutral opinion will not take these measures.

In earlier modelling work, sustained circulation of the health opinions from both sides of the spectrum required the presence of an outbreak (2, 4, 23). However, frequently, the opinions persist without the disease being present. In this case, the opinion switching rates depend on the number/density of the carriers of these opinions. Another important consideration is the functional definition of the opinion switching rate. Often it is captured by a mass action term (2, 4, 14, 18) that may not necessarily reflect the reality. We address both of these considerations. In our model, individuals switch between opinions as a result of communication with individuals of the opposing opinion, with a switch rate that is a positive non-decreasing function of the density of individuals holding the opposing opinion. Here we consider a broader family of functions to describe the rate of switching, which includes linear, saturating, and sigmoidal functions. We couple opinion dynamics with an epidemic model by allowing the rate of switching to the health-positive opinion to depend on the disease prevalence. With respiratory diseases as influenza or COVID-19 in mind, we consider a population that mixes assortatively by opinions.

Using bifurcation and stability analysis, we investigate the opinion distribution landscape in the absence of disease. The dynamics in a disease-free state both highlight the key considerations for the design of information intervention prior to the outbreak, as well as set the stage for epidemic dynamics in case an infectious disease enters the population. We analyze for which distributions of opinions in the population an outbreak of an infectious disease can occur, i.e. how the distribution of opinions impacts the basic reproduction number of the infection. We then explore the coupled opinion-epidemic dynamics using numerical bifurcation analysis. Finally, we describe parameter regions, for which damped/sustained oscillatory dynamics may appear, and give conditions under which a disease can be eradicated even when the basic reproduction number is above 1.

## Results

### A model for competing opinions

In the context of an infectious disease, we consider a scenario where two relevant mutually exclusive health opinions, *a* and *b*, circulate in a population. We denote with *a* a health-positive opinion whereupon an individual holding it adapts measures that reduce the probability of contracting the disease, and *b* denotes a health-neutral opinion such that its holder does not modify their behavior. Thus, the population is split into individuals who believe *a, N*_*a*_ and those who believe *b, N*_*b*_. In this work we use word “density” to denote a proportion of total population. Thus, the proportion of population (density) that holds opinion *a* is denoted by *n*_*a*_, while the density of population *N*_*b*_ is *n*_*b*_. We assume that individuals regardless of their opinion have

on average *c* social contacts per week. We use the term “social contacts” to denote interactions that may lead to switching of opinions. Additionally, we consider the possibility of assortative preference to mix with individuals of the same opinion. The degree of assortative mixing is denoted by *ω*, 0 ≤ *ω* ≤ 1, with *ω* equal to 0 describing the situation where individuals interact without regard about the opinion held (fully proportionate mixing) and *ω* equal to 1 denotes fully assortative mixing where individuals only mix with individuals which share their opinion. For 0 ≤ *ω* ≤ 1, *ω* indicates the proportion of contacts that occur only with individuals sharing the same opinion, while 1 − *ω* fraction of contacts occur with holders of each opinion, proportionate to the density of respective population.

Individuals 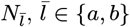 may change their opinion upon contact with individuals with the opposing opinion, 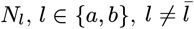. The rate of switching is described by a density-dependent function *f*_*l*_(*n*_*l*_), multiplied by social contact rate *c*, and the likelihood of mixing with individuals regardless of their opinion, 1 − *ω*. We assume the switch rate functions *f*_*l*_(*n*_*l*_) to be positive, continuous and increasing, and define

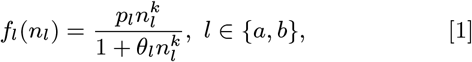

where *p*, 0 ≤ *p* ≤ 1 is the per contact probability of switching from opinion 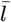 to opinion *l*. Parameters 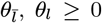, and *k, k* ≥ 1 specify the shape of the response function. Observe that the switch rate to an opinion is zero, if there are no individuals with that opinion in the population.

Three types of response functions can be distinguished depending on parameters *k* and *θ* (Figure 1**a**): (1) for *k* = 1 and *θ*_*a*_ = *θ*_*b*_ = 0 the switch rate function is linear; (2) for *k* = 1 and *θ*_*a*_, *θ*_*b*_ *>* 0 the switch rate function is saturating for large densities; (3) for *k >* 1 and *θ*_*a*_, *θ*_*b*_ *>* 0 the switch rate function is sigmoidal. In ecology, very similar functions have been derived from first principles to describe the functional response of predator population density to the density of available prey, and are known as Holling type I, II, III functional response (24).

**Fig. 1.**
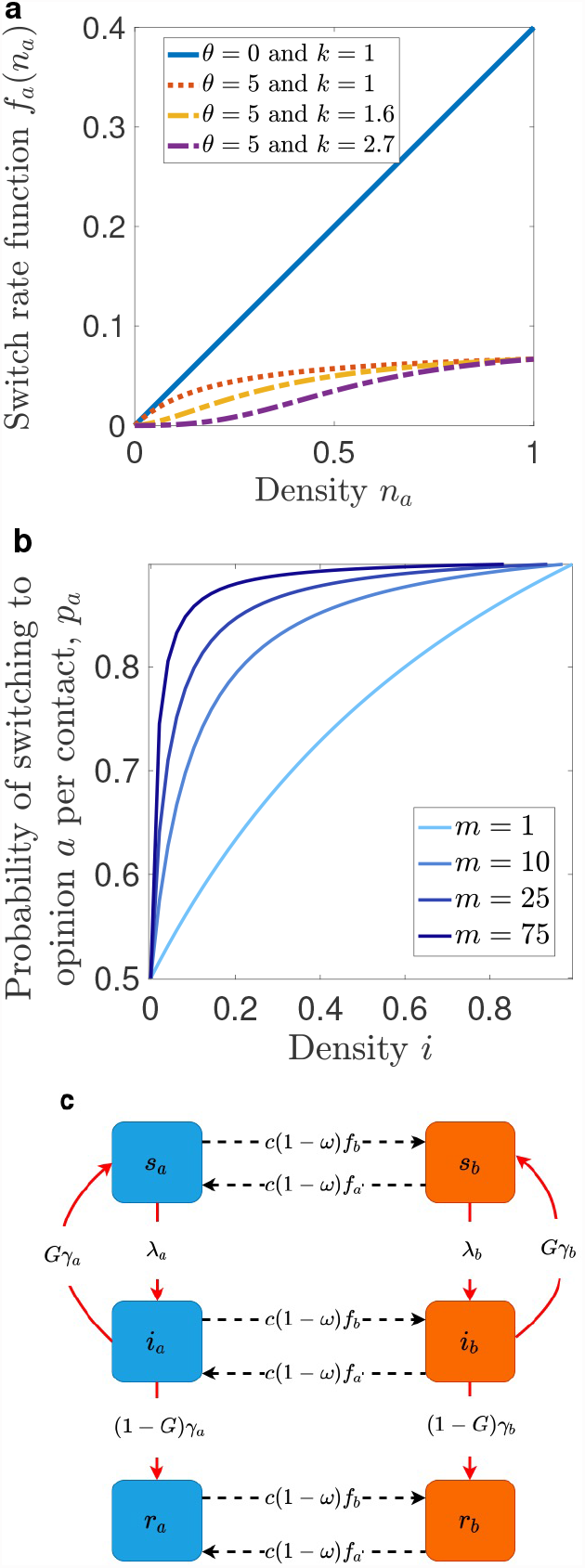
Coupling of opinion dynamics and infection transmission dynamics. **a** Switch rate function to opinion *a* depending on density *n*_*a*_. For *θ* = 0 and *k* = 1 the switch rate is linear (blue); for *θ >* 0 and *k* = 1 the switch rate is saturating (red); for *θ >* 0 and *k >* 1 the switch rate is sigmoid (yellow and violet). **b** Per contact probability of switching to opinion *a* for different values of *m* as a function of the density of infected individuals, *i*. **c** Flowchart of coupled opinion and infection transmission model for two types of infectious diseases: SIS model (*G* = 1) and SIR model (*G* = 0); black dashed arrows denote opinion transitions, red solid arrows denote epidemiological transitions.

In this work we investigate long term opinion dynamics for each one of these response functions. However, note that, to describe the diffusion of innovations or opinions in a population, sigmoidal functions have been used (25). These functions capture the trend whereupon the spread of an opinion *l* is very slow as long as only a small proportion of the population holds this opinion, and slows down again when the proportion of the population *N*_*l*_ is large, with fast growth in between. The saturation for high density of *N*_*l*_ mimics the saturation of information effect, whereupon the information loses its impact once it has been received several times. In our model, both opinions spread according to a sigmoidal response function, possibly with different shapes. This leads to a system in which opinions compete and may either co-exist or drive each other to extinction.

We assume that opinion dynamics are fast compared to the natural demographic processes, and therefore do not include demographic processes in the model.

### A model coupling opinion dynamics and epidemic dynamics

We consider a disease that follows a Susceptible-Infected-Recovered (SIR) or a Susceptible-Infected-Susceptible (SIS) model. To investigate the effect of feedback between disease dynamics and opinion dynamics on the course of an epidemic, we couple the above described framework of opinion competition with a SIR or SIS infection transmission model (Figure 1**c**). For both types of disease dynamics, individuals become infected and infectious at rate *λ*, which depends on the prevalence of infection, *i*. Infectious individuals recover with rate *γ*, either becoming susceptible again (SIS model) or becoming immune (SIR model).

Each individual has an opinion and an infection status. We denote the density of susceptible individuals holding opinion *a* with *s*_*a*_, the density of infectious individuals holding the same opinion with *i*_*a*_, and the density of recovered individuals with *r*_*a*_. Similarly, *s*_*b*_, *i*_*b*_, and *r*_*b*_ denote the densities of individuals who hold opinion *b* in the respective epidemiological states. Individuals *N*_*a*_ have a lower probability of acquiring infection than individuals *N*_*b*_, i.e. *β*_*a*_ ≤ *β*_*b*_. We assume that the measures taken by *N*_*a*_ only reduce their susceptibility, and that infectivity and the recovery rate are the same for the two types of individuals. Note that the parameters *β*_*a*_, *β*_*b*_ implicitly include the transmission-relevant contact rate, which may differ from the social contact rate *c*. Finally, we consider the case where assortativity also applies to infection-relevant contacts, such that in terms of physical contacts, the individuals can prefer to mix with individuals who have the same health opinion. Therefore the rates with which individuals *s*_*a*_ and *s*_*b*_ are specified by the following equations:

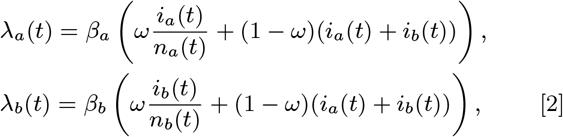

The infection status of individuals does not modify the rate with which they switch their opinion. However, infection spread in the population can affect opinion dynamics. Here, we consider the case of individuals obtaining information about disease spread that is available publicly via media and health authorities. In our model, with increasing prevalence of infection *i* = *i*_*a*_ + *i*_*b*_, opinion *a* gains in popularity, which is represented by an increase in the probability of switch to opinion *a* per contact, *p*_*a*_. We assume that

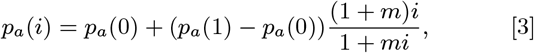

where *p*_*a*_(0) is the switching rate per contact in the disease-free state, and *p*_*a*_(1) is the switching rate when the entire population is infected; *m* is a constant that determines how fast *p*_*a*_ increases with increasing prevalence (see Figure 1**b**). Thus, as prevalence of infection increases, so does the switch rate to opinion *a* (Eq. (1)). Probability of switching to opinion *b* per contact, *p*_*b*_, remains fixed throughout the outbreak.

The dynamics are described by a flow diagram shown in Figure 1**c** and are captured by system of ordinary differential equations (6) in Methods section.

Model parameters are summarized in Table 1. In numerical analysis, we use the indicated parameter values, unless stated otherwise. We give the justification for the selection of the values later in the text.

**Table 1.**
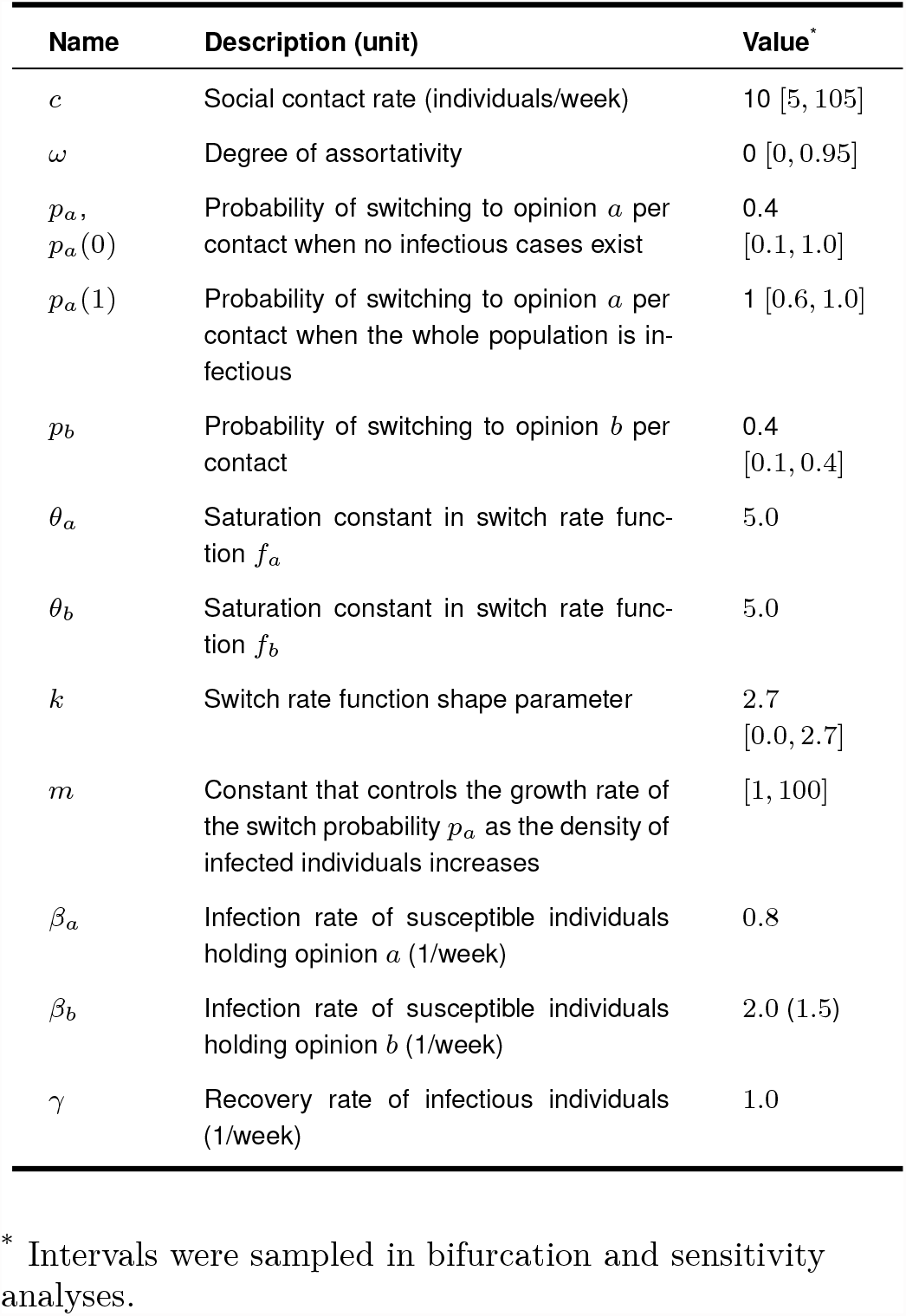
Summary of model parameters described by system (6) and ranges of values used in numerical examples.

| Name | Description (unit) | Value* |
| --- | --- | --- |
| $c$ | Social contact rate (individuals/week) | 10 [5, 105] |
| $\omega$ | Degree of assortativity | 0 [0, 0.95] |
| $p_a, p_a(0)$ | Probability of switching to opinion $a$ per contact when no infectious cases exist | 0.4 [0.1, 1.0] |
| $p_a(1)$ | Probability of switching to opinion $a$ per contact when the whole population is infectious | 1 [0.6, 1.0] |
| $p_b$ | Probability of switching to opinion $b$ per contact | 0.4 [0.1, 0.4] |
| $\theta_a$ | Saturation constant in switch rate function $f_a$ | 5.0 |
| $\theta_b$ | Saturation constant in switch rate function $f_b$ | 5.0 |
| $k$ | Switch rate function shape parameter | 2.7 [0.0, 2.7] |
| $m$ | Constant that controls the growth rate of the switch probability $p_a$ as the density of infected individuals increases | [1, 100] |
| $\beta_a$ | Infection rate of susceptible individuals holding opinion $a$ (1/week) | 0.8 |
| $\beta_b$ | Infection rate of susceptible individuals holding opinion $b$ (1/week) | 2.0 (1.5) |
| $\gamma$ | Recovery rate of infectious individuals (1/week) | 1.0 |

\* Intervals were sampled in bifurcation and sensitivity analyses.

To calculate the basic reproduction number *R*_0_ for this model, we used the Next Generation Matrix method described in (26).

Then *R*_0_ is given by the spectral radius of matrix *FV* ^*−*1^ with

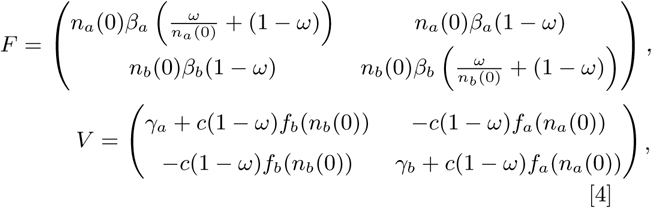

Here (*n*_*a*_(0), *n*_*b*_(0)) are given by the opinion distribution at the start of the outbreak and depend on *k, θ*_*a*_, *θ*_*b*_, *p*_*a*_*/p*_*b*_.

For a population, in which only one of the two opinions is present (“monobelief” population), the epidemic dynamics are reduced to the basic SIS/SIR dynamics with a basic reproduction number that is determined by the parameters of the dominating opinion:

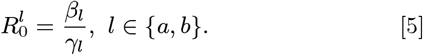

### Dynamics of competing opinions

To understand the effect of the coupling between the disease spread and opinion competition on infection transmission, we first need to consider the dynamics of opinions in the disease-free population.

The model indicates that when either one of the two opinions dominates the population (“monobelief” population), then this remains unchanged until individuals of the opposing opinion, enter the population from outside. As we are mainly interested in situations where two opinions compete in the population, we investigated for which parameter regions a stable co-existence of two opinions is possible. This co-existence depends on the shape of the switch rate functions, *f*_*l*_, *l* ∈ {*a, b*}, but not on the social contact rate *c* or the assortativity parameter *ω*, as these are assumed to be the same for both opinions (Supporting information (SI), Supplementary text).

For linear switch rate functions (Eq. (1), *θ*_*l*_ = 0, *k* = 1), the stable co-existence of opinions is not possible (Figure 2). As there is no density dependence, the growth of the switch rate functions does not slow down even when the majority of the population is following a certain opinion. If initially both opinions are present, the opinion with the larger probability of switching per contact *p*_*l*_ will take over. If, for example, opinion *a* is introduced into a monobelief population with opinion *b*, it will only be able to persist if *p*_*a*_ is larger than *p*_*b*_. In this case, the population will eventually switch to opinion *a* regardless of the initial density of individuals who hold it (Figure 2**b**). Assortativity degree (*ω*) and contact rate (*c*) while not affecting the outcome of the opinion competition, determine the speed with which the mono-belief state is reached, such that higher assortativity and lower contact rate prolong the transient period.

**Fig. 2.**
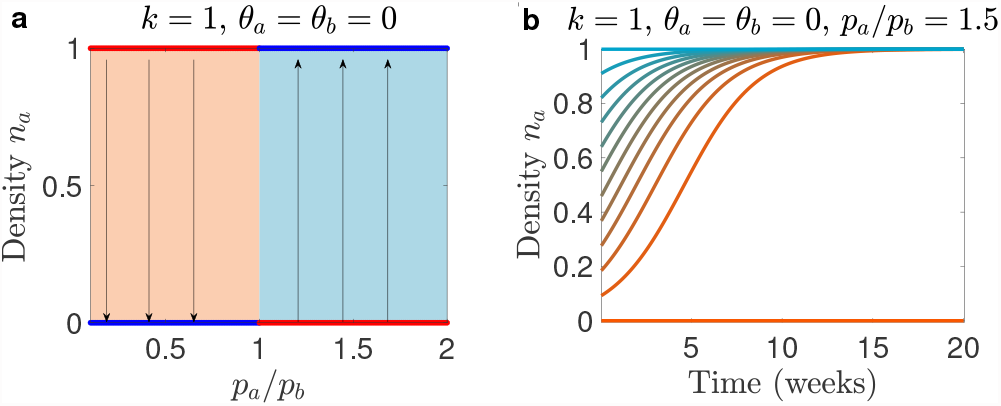
Opinion competition dynamics for a linear switch rate function. We consider opinion dynamics in the disease-free population. **a** Bifurcation diagram of *n*_*a*_ as a function of *p*_*a*_*/p*_*b*_. Red lines: unstable equilibria, blue lines: stable equilibria. Orange area: basin of attraction of the mono-belief *b* equilibrium, blue area: basin of attraction of the mono-belief *a* equilibrium. **b** Temporal dynamics of *n*_*a*_ for *p*_*a*_*/p*_*b*_ = 1.5. All solutions where initially both opinions are present converge to the state where opinion *a* dominates.

If the switch rate functions are non-linear (Eq. (1), *θ*_*l*_ *>* 0, *k* ≥ 1) the opinions can co-exist in a steady state (Figure 3). For switch rate functions that are saturating but not sigmoidal (*θ*_*l*_ *>* 0, *k* = 1), either stable co-existence is possible, or one of the mono-belief solutions is stable. It depends on the two switch rate functions, whether co-existence is possible or not (Figures 3**a**, 3**d**, 3**g**). Stable co-existence of opinions is possible in the case when the switching functions exhibit saturation at high density of an opinion. Subsequently, the growth of the switch rate function for the dominant opinion slows down when the majority of the population is following that opinion. The stable co-existence state is attracting for all initial situations, in which both opinions are present. The distribution of opinions at this steady state depends entirely on the ratio *p*_*a*_*/p*_*b*_ and not on *p*_*a*_ and *p*_*b*_ separately (SI, Supplementary text). The larger the ratio *p*_*a*_*/p*_*b*_, the higher is the equilibrium density of *N*_*a*_ individuals. If permanent co-existence of opinions is impossible, the opinion with higher switch rate per contact (*p*_*l*_, *l* ∈ {*a, b*}) will take over the population. The interval of *p*_*a*_*/p*_*b*_, in which opinions can co-exist, depends on the saturation constants of the switch rate functions, *θ*_*l*_, *l* ∈ {*a, b*}. The higher these are (i.e. the faster saturation is achieved) the wider is the *p*_*a*_*/p*_*b*_ interval, in which opinions can co-exist. Intuitively, the faster the switch rate functions become saturated, the larger differences between the probabilities of switching per contact can be while still allowing stable co-existence of opinions. For mathematical derivations and further elaborations, see SI, Supplementary text.

**Fig. 3.**
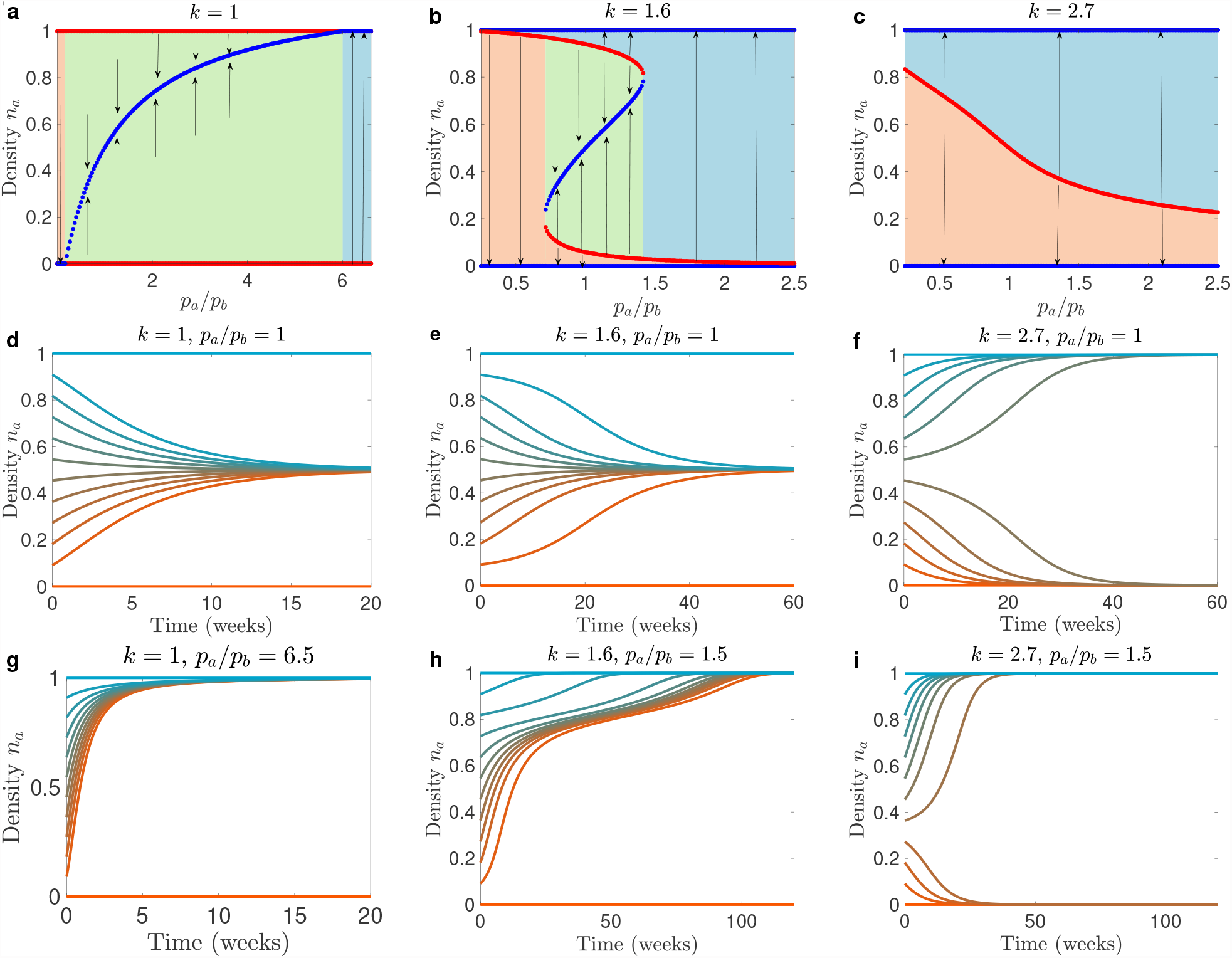
Opinion competition dynamics for saturating and sigmoidal switch rate functions. The upper row shows bifurcation diagrams of *n*_*a*_ as a function of *p*_*a*_*/p*_*b*_. For **a** a saturating switch rate function; **b** and **c** sigmoidal switch rate functions. Red lines: unstable equilibria, blue lines: stable equilibria. Orange area: basin of attraction for the equilibrium with opinion *b* dominating; blue area: basin of attraction for the equilibrium where opinion *a* dominates; green area: basin of attraction of a stable co-existence equilibrium. **d**-**i** Temporal dynamics of *n*_*a*_ for different switch rate functions and ratios *p*_*a*_*/p*_*b*_. In all panels *θ*_*a*_ = *θ*_*b*_ = 5.

If the switch rate functions are sigmoidal (Eq. (1), *θ*_*l*_ *>* 0, *k >* 0), at least one stable co-existence state of opinions is possible for some parameter regions (Figure 3**b**, 3**e**). Additionally, monobelief population states are always locally attracting. I.e., if, for example, the population starts with a sufficiently large majority believing opinion *a*, then after some time the entire population will hold this opinion.

If for a given set of parameters there is only a single unstable co-existence equilibrium (Figures 3**b** and 3**c**), the population always ends up as a monobelief population, but it depends on the initial distribution of opinions to which mono-belief it will converge. The proportions of *n*_*a*_ and *n*_*b*_ at this unstable steady state depend on the ratio *p*_*a*_*/p*_*b*_. The higher this ratio, the lower is *n*_*a*_. This unstable equilibrium separates the state space into the basins of attraction of the *a*-monobelief and *b*-monobelief populations. This implies that the population with the higher associated switch probability per contact *p*_*l*_ requires a smaller proportion of individuals of that opinion to invade. This is illustrated in Figure 3**g**), where *p*_*a*_ is 1.5 times higher than *p*_*a*_, hence it requires much fewer individuals of opinion *a* to take over the population.

If, on the other hand, for a given fixed set of parameters several steady states are possible, then their number is odd and at least one of them is locally attractive. For the interpretation of the model, only locally stable steady states are of interest as states in which two opinions can co-exist. Unstable steady states are relevant as boundaries between basins of attraction. In our numerical experiments, we observed at most three different steady states, one of them a stable co-existence state (see Figure 3**b**). Our analysis and numerical experiments indicate that existence of a stable co-existence state of opinions depends on values of *p*_*a*_*/p*_*b*_, *θ*_*a*_, *θ*_*b*_, and *k* (SI, Supplementary text).

If there are three steady states, two of them are repelling and one is attracting, such that the density *n*_*a*_ for the attracting state is between the densities *n*_*a*_ for the repelling states. Therefore, the repelling states mark the boundaries of the basins of attraction for the attracting states. From the bifurcation diagram (Figure 3**b**) we observe that there are two points where the dynamics of the system change as *p*_*a*_*/p*_*b*_ increases from zero (left and right edges of the green region on Figure 3**b**). These are saddle node bifurcation points which mark the appearance and disappearance of a pair of steady states. If *p*_*a*_*/p*_*b*_ is to the left of the green region, then in order to take over the population, nearly the whole population should hold opinion *a*. Stable co-existence of opinions is impossible. As *p*_*a*_*/p*_*b*_ increases and passes the left edge of the green region, this proportion *n*_*a*_ needed for opinion *a* to take over the population declines (Figure 3**b**, upper red curve in the green region). More importantly, stable co-existence with opinion *b* is now possible and requires a much smaller initial proportion of *n*_*a*_ for persistence of *a*. (Figure 3**b**, lower red curve in the green region). As *p*_*a*_*/p*_*b*_ increases past the right edge of the green region, the “invasion” density threshold for opinion *a* further declines. Moreover, as stable co-existence is not possible anymore, it becomes the threshold for complete taking over of the population by opinion *a*.

### Epidemic dynamics in a population with competing opinions

For the purposes of analysis of the feedback between opinion competition and infection dynamics, we are mainly interested in the situation where health-positive and health-neutral opinions can co-exist in a steady state and the monobelief population steady states are locally stable. We therefore focus our attention on sigmoidal opinion switch rate functions and on the parameter region where stable co-existence of opinions is possible. We assume that an infectious disease invades a population, in which the two opinions co-exist at the stable steady state.

The opinion switch rate-related parameters are fixed at *k* = 1.6, *θ*_*a*_ = *θ*_*b*_ = 5. Thus the switch rates for both opinions are sigmoidal functions. We fix *p*_*b*_ = 0.4. For most of the simulations *p*_*a*_ and *p*_*a*_(0) are fixed to 0.4, thus *p*_*a*_*/p*_*b*_ = 1 and the stable co-existence of opinion equilibrium has 50/50 distribution of health-positive and health-neutral individuals. Probability of switching to to opinion *a* per contact when the whole population is infectious *p*_*a*_(1) is bounded by the largest possible value it can have, 1. Assortativity degree *ω* and social contact rates *c* are varied on the intervals which are sufficiently wide to recover full range of qualitative dynamics.

We consider the dynamics of a respiratory non-fatal infectious disease similar to flu. We assume that the infectious period lasts on average a week, thus we fixed *γ*_*a*_ = *γ*_*b*_ = 1 per week. Furthermore, we assume that in a population where opinion *a* is dominant, the infection cannot spread because the health-positive opinion leads to protective behavior that prevents an outbreak of the infection. In a population, where opinion *b* dominates, this health-neutral opinion enables the infection to spread. The transmission parameters are set as follows. The infection rate of susceptible individuals holding the health-positive opinion *a* is fixed *β*_*a*_ = 0.8 per week, and the infection rate for individuals holding the health-positive opinion *b* is fixed *β*_*b*_ = 2 per week for SIR model and *β*_*b*_ = 1.5 per week for SIS model. This difference of values was necessary, since in the case of SIS the pool of susceptible individuals is being constantly replenished. These settings imply that 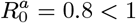 and 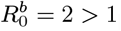.

### Basic reproduction number

In a situation where both opinions are present at the time the infectious disease comes into the population, the basic reproduction number *R*_0_ depends on the proportions *n*_*a*_ and *n*_*b*_. We assume that these proportions are at steady state at the moment of onset of an epidemic. Recall that *c* and *ω* do not influence this steady state distribution of opinions, so the initial situation is the same for all values of those parameters. We therefore can investigate how social contact rate *c* and degree of assortativity *ω* impact the epidemic dynamics without changing the initial steady state of the system. By varying *c* and *ω*, we change the way the population can adapt to an emerging outbreak by communicating about health-positive behavior. With increasing *c*, opinions can spread faster, while with increasing *ω*, opinions are more restricted to their subpopulation.

In Figure 4 we investigated how the basic reproduction number *R*_0_ changes with changing social contact rate *c* and assortativity degree *ω* for three settings of the ratio *p*_*a*_*/p*_*b*_: 0.8, 1, and 1.25.

**Fig. 4.**
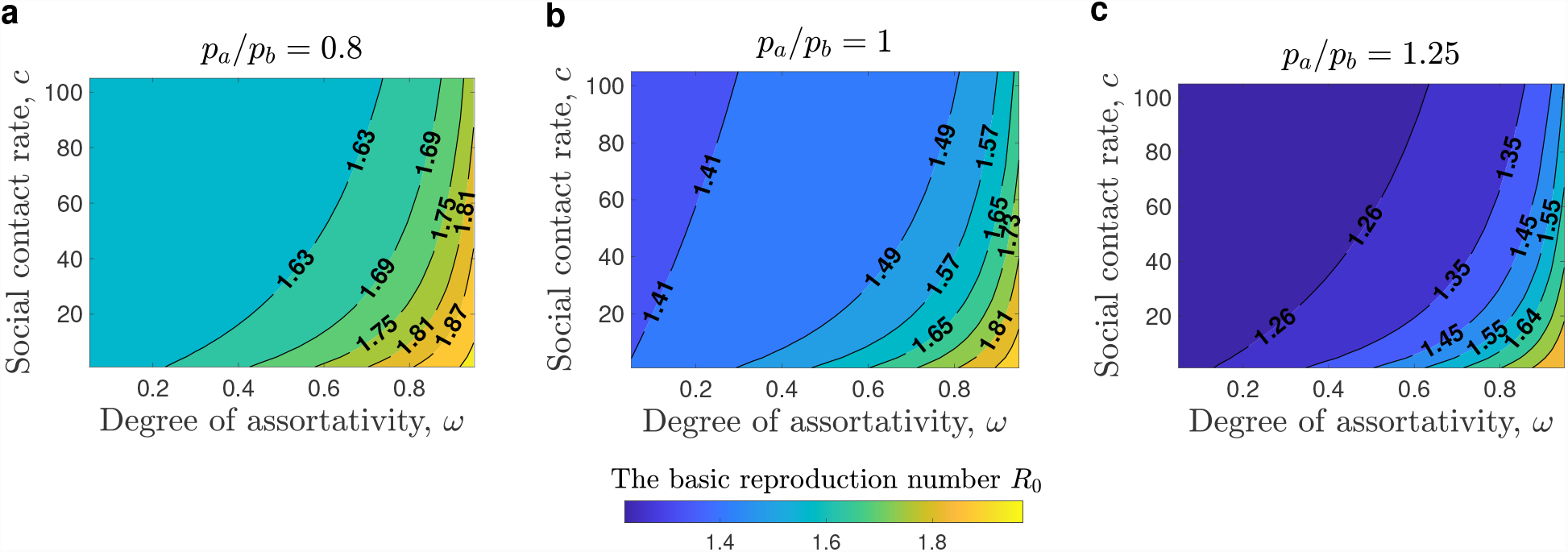
Impact of mixing patterns on basic reproduction number *R*_0_. **a, b**, and **c** show contour maps of *R*_0_ as a function of the social contact rate *c* and the assortativity *ω*. **a** For *p*_*a*_*/p*_*b*_ = 0.8 the initial distribution of opinions is (*s*_*a*_(0), *s*_*b*_(0)) = (0.35, 0.65). **b** For *p*_*a*_*/p*_*b*_ = 1 we have (*s*_*a*_(0), *s*_*b*_(0)) = (0.5, 0.5). **c** For *p*_*a*_*/p*_*b*_ = 1.25 we have (*s*_*a*_(0), *s*_*b*_(0)) = (0.65, 0.35). The infection rate of susceptible individuals holding opinion *a* is fixed *β*_*b*_ = 2, the value used to investigate the dynamics for the SIR system. For the same set of figures with *β*_*b*_ = 1.5, the value used to investigate the dynamics of the SIS model, see Figure S2 in SI.

For all three settings of ratio *p*_*a*_*/p*_*b*_, the basic reproduction number increases as assortativity *ω* increases, and decreases as the social contact rate (*c*) increases. As the ratio *p*_*a*_*/p*_*b*_ increases, the basic reproduction number decreases. We note that for high assortativity, the effect of increasing *c* is smaller than for low assortativity. Overall, we conclude that increasing assortativity slows down the spread of opinions and therefore leads to higher values of *R*_0_. Conversely, increasing social contact rate *c* leads to faster opinion spread and therefore to lower *R*_0_. Therefore, strong assortative mixing by opinions can facilitate the outbreak of an infectious disease.

### SIR model with opinion competition

In this section, we consider the dynamics beyond the start of an outbreak for an SIR-type disease and investigate how it depends on *c* and *ω*. We fixed *p*_*a*_(0)*/p*_*b*_ = 1 and *p*_*a*_(0)*/p*_*b*_ = 2.5 and used the respective stable co-existence distribution (*n*_*a*_ = 0.5, *n*_*b*_ = 0.5) as the initial state of the population. We seeded infection by setting *i*_*b*_(0) = 6 ×10^*−*8^ and *s*_*b*_(0) = *n*_*b*_(0) − *i*_*b*_(0).

We investigated the effect of the feedback between opinion competition and infection dynamics on the epidemic peak and on the peak density of the *N*_*a*_ population during and after the outbreak. We used three settings for parameter *m*, which affects the sensitivity of the population to the growth in prevalence of infection. As the prevalence increases, *p*(*a*) now increases, and this can be slower (*m* = 25) or faster (*m* = 75) (Figure 5).

**Fig. 5.**
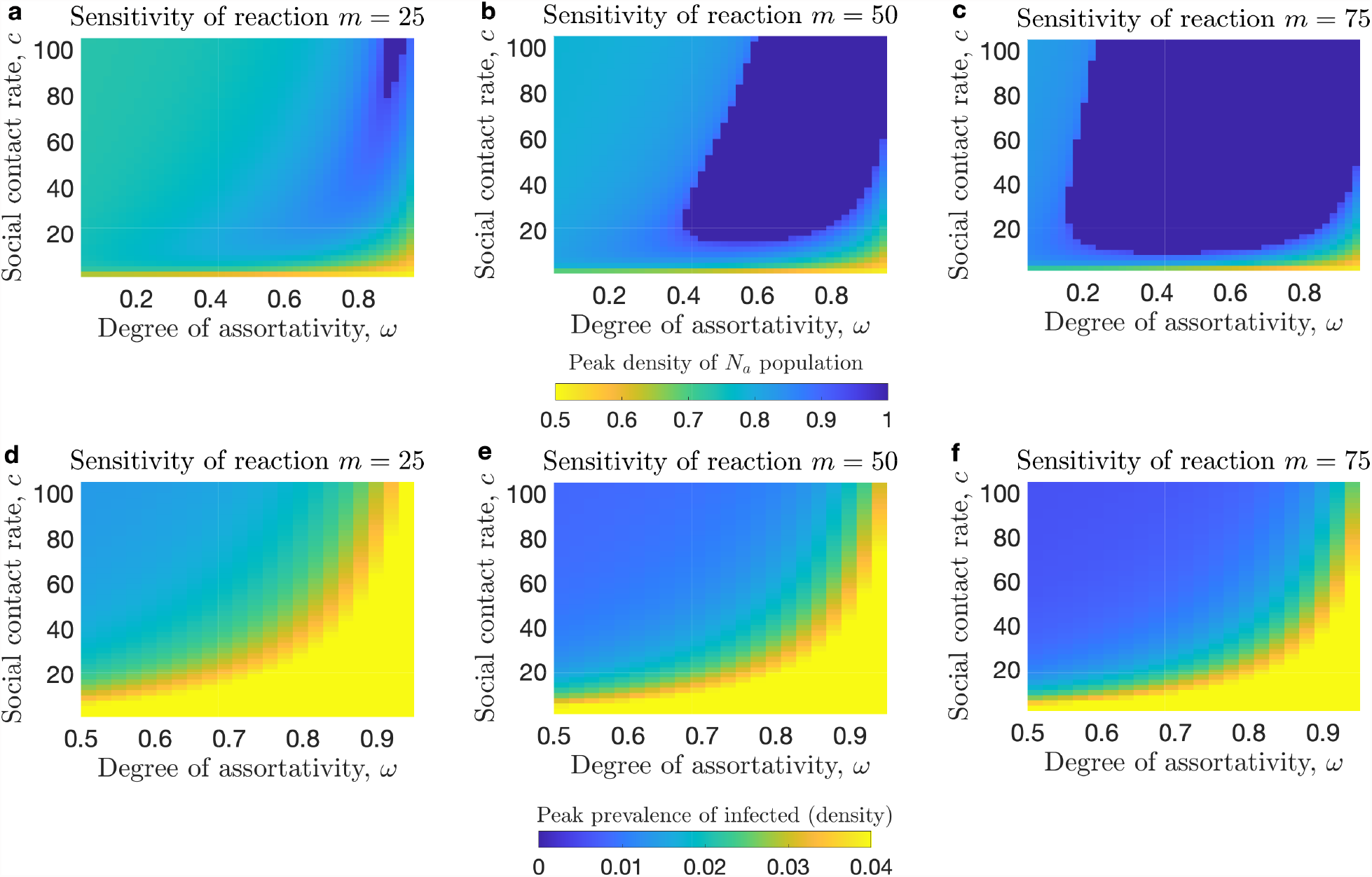
Impact of social contact rate and assortativity on epidemic dynamics. We consider the dynamics of the SIR system for three scenarios for the sensitivity of the population to increasing prevalence of infection as denoted by parameter *m, m* = 25 for **a** and **d**, *m* = 50 for **b** and **e**, and *m* = 75 for **c** and **f. a, b**, and **c** show heat maps of the peak density of the *N*_*a*_ population; in the dark blue region the population converts to opinion *a*. **d, e**, and **f** show contour maps of the peak prevalence.

For all three scenarios the peak prevalence is higher for lower contact rates and higher assortativity. The higher is the sensitivity of the population *m*, the lower is the prevalence peak.

In Figure 6, the temporal dynamics are shown for some parameter combinations. As a consequence of the feedback between the disease and infection dynamics, the density of individuals who hold opinion *a* temporarily increases, with eventual return of the population to the pre-outbreak opinion distribution. However, for some parameters settings, the population may convert completely to opinion *a*, thereby preserving the memory of the past outbreak. We investigated the parametric region, in which this conversion to *a* occurs (Figure 5 and Figure S5 in SI). From Figure **??** it follows that high sensitivity of the population to rise in prevalence of infection, as reflected in parameters *p*_*a*_(1) and *m* and a high social contact enable conversion to opinion *a*. In addition, a high degree of assortativity also enables opinion *a* to become dominant (dark blue region in Figures 5**a**, 5**b**, and 5**c** and in Figures S5**a** and S5**b** in SI). This is unexpected, since high assortativity slows down opinion exchange. However, since high assortativity also leads to a large *R*_0_, it leads to a rise in prevalence, and therefore increases the probability of switching to opinion *a*. More technically, the convergence of the population to a monobelief *a* population requires that the *n*_*a*_ component crosses into the basin of attraction of the mono-belief *a* steady state (red lines on Figures 6**a** and 6**d**). Several conditions make this event possible: (1) a high prevalence of infection, (2) a fast response of the population to increasing prevalence; (3) a high rate of switching from opinion *b* to *a*.

**Fig. 6.**
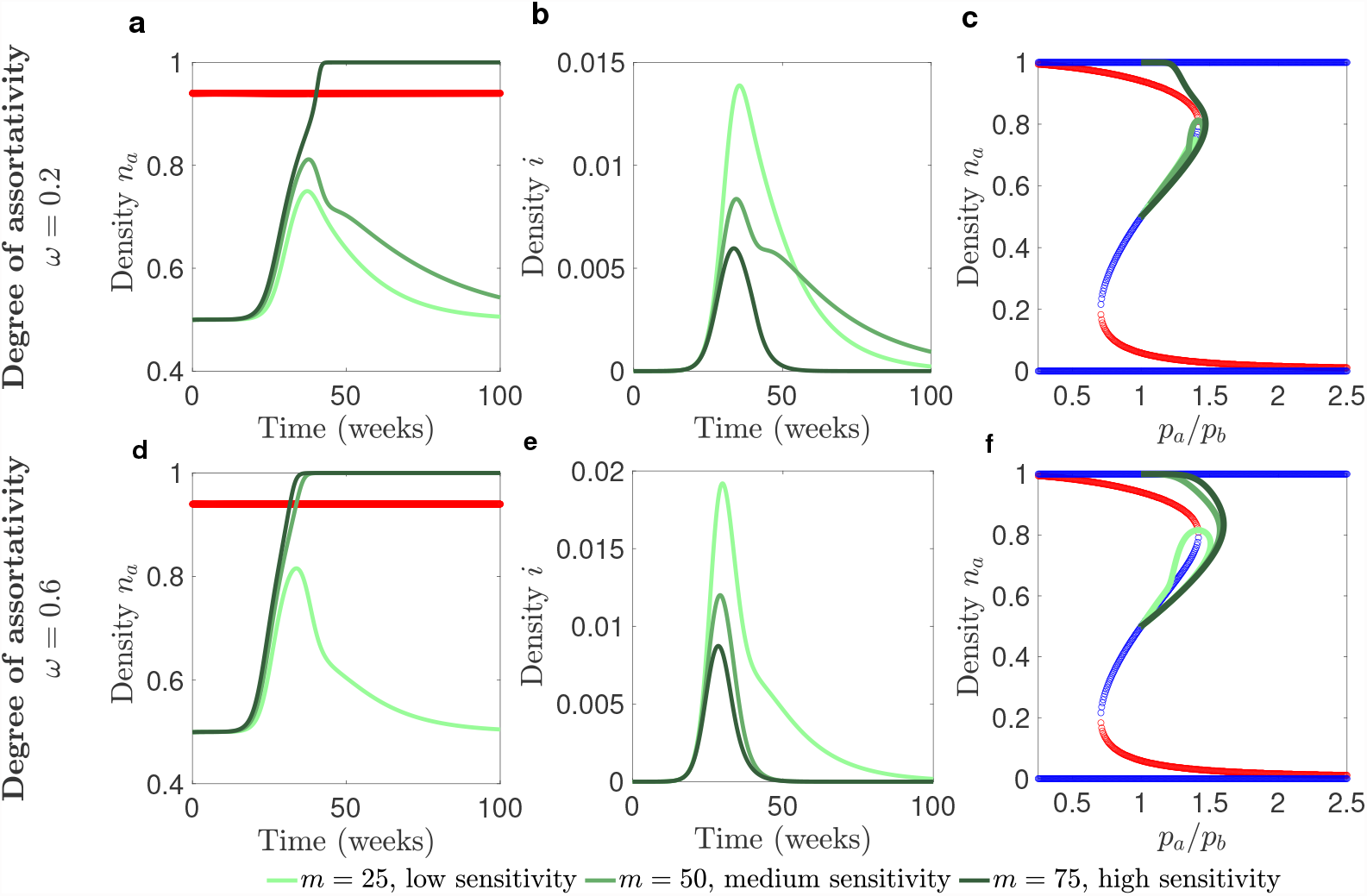
Impact of assortativity on the population adopting opinion *a*. We consider the dynamics of the SIR system. **a** and **d** show density of *N*_*a*_ population *n*_*a*_ in time, **b** and **e** show the prevalence of infectious cases in time, **c** and **f** show phase diagrams for three different solutions overlaid over a bifurcation diagram for density of *N*_*a*_ population, *n*_*a*_, component of permanent distributions. **a, b**, and **c** are plotted for degree of assortativity *ω* = 0.2, **a, b**, and **c** - for *ω* = 0.6. Social contact rate is fixed *c* = 40.

In contrast with the standard SIR epidemic, whose dynamics display a single peak only, in a situation with feedback between the disease dynamics and opinions dynamics multiple epidemic peaks may appear (Figure 7). Our numerical analyses indicate that in order for multiple epidemic peaks to appear there should be a pronounced difference between population *N*_*a*_ and *N*_*b*_ in terms of the preventative measures they adapt (as reflected in parameters *β*_*a*_ and *β*_*b*_). The upper boundary of the region in *β*_*a*_ − *β*_*b*_ subspace where multiple peaks appear marks the region where the population switches to opinion *a* (red curve). Therefore, for a fixed *β*_*b*_ as *β*_*a*_ increases multiple peaks appear as the population moves to the *a*-monobelief state (Figure 8). The number of peaks grows as *β*_*a*_ moves closer to the boundary. Note that in our analyses, we considered a local maximum of prevalence to be a peak if it exceeded 10^*−*8^. Moreover, the more sensitive the population is to increases in the prevalence of infection (as reflected by parameter *m*), the larger is the number of peaks that will appear in the region adjacent to the boundary where switch of the population to opinion *a* occurs, see Figure 7 and Figure S4 in SI. Finally, if the probability of switch to opinion *a* in the population without infection, *p*_*a*_(0) is significantly smaller than the probability of switch to opinion *b, p*_*b*_ the region in *β*_*a*_ − *β*_*b*_ space where multiple peaks exist is larger (see Figure S3 in SI).

**Fig. 7.**
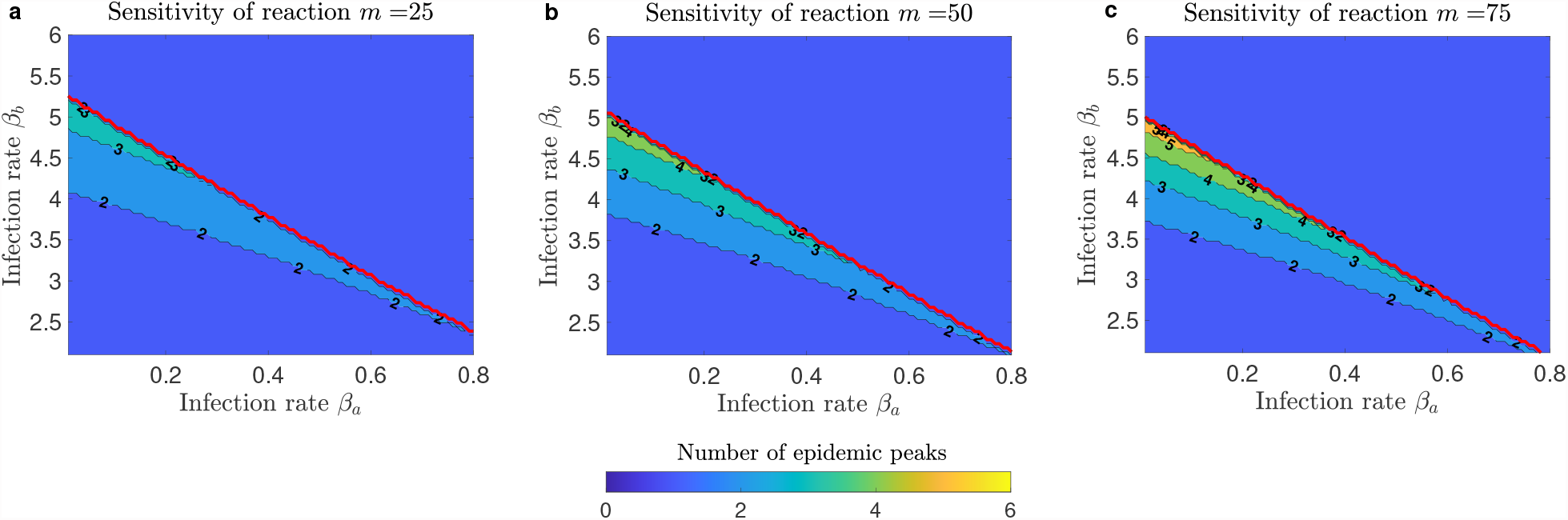
Regions of multiple epidemic peaks resulting from feedback between disease dynamics and opinion dynamics. We consider the dynamics of the SIR model. **a, b**, and **c** are contour plots of the number of prevalence peaks for different values of infection rates *β*_*a*_ and *β*_*b*_ for different sensitivity *m* of the population to increasing prevalence: **a** *m* = 25, **b** *m* = 50, and **c** *m* = 75. The social contact rate is fixed at *c* = 40, and the probability of switch to opinion *a* per contact when the entire population is infected is fixed *p*_*a*_ (1) = 1, and the assortativity degree is fixed *ω* = 0. The area above the red curve denotes the outcome where the population switched to opinion *a*. As *m* increases this region expands.

**Fig. 8.**
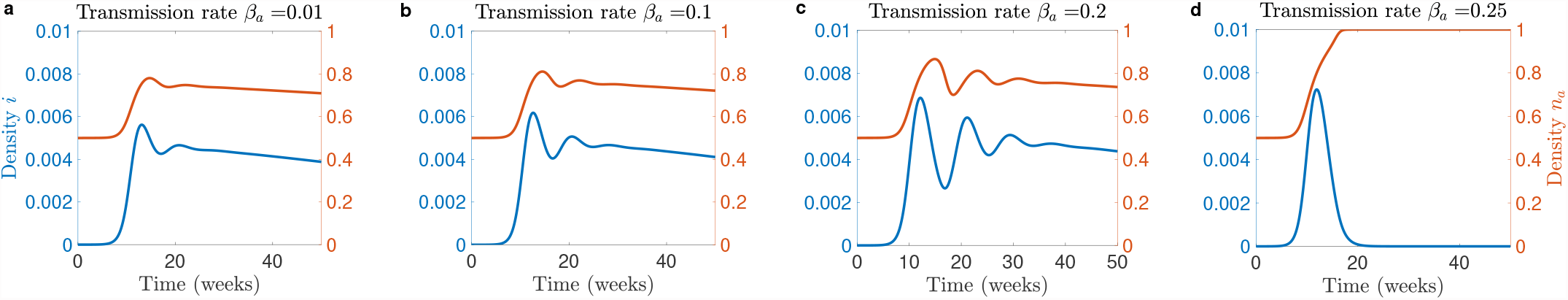
Temporal dynamics with multiple epidemic peaks resulting from feedback between disease dynamics and opinion dynamics. We consider the dynamics of the SIR system. Panels show time series of infection prevalence, and of the density of the *N*_*a*_ population, *n*_*a*_ for different values of infection rate *β*_*a*_. The social contact rate was fixed at *c* = 40, the upper bound of the probability of switching to opinion *a* was set to *p*_*a*_ (1) = 1, the sensitivity parameter *m* was set to *m* = 75, the infection rate of *N*_*b*_ individuals was set to *β*_*b*_ = 4.15, and the assortativity degree is fixed *ω* = 0.

In summary, for SIR-dynamics we find that feedback between opinion dynamics and epidemic dynamics can substantially change the epidemic outcomes. The basic reproduction number *R*_0_ and the peak of an outbreak can be higher if there is assortative mixing by opinion. In addition, multiple epidemic peaks can occur and the response to an epidemic can lead to a shift of the population to a state, in which only the health-positive opinion is circulating.

### SIS model with opinion competition

Similarly, for a SIS-infection, coupling between opinion competition and disease dynamics can lead to opinion *a* taking over the population (Figure 9), and to the appearance of oscillatory epidemic dynamics (Figure 10). For the SIS epidemic, these oscillatory dynamics can be sustained epidemic cycles instead of damped oscillations.

**Fig. 9.**
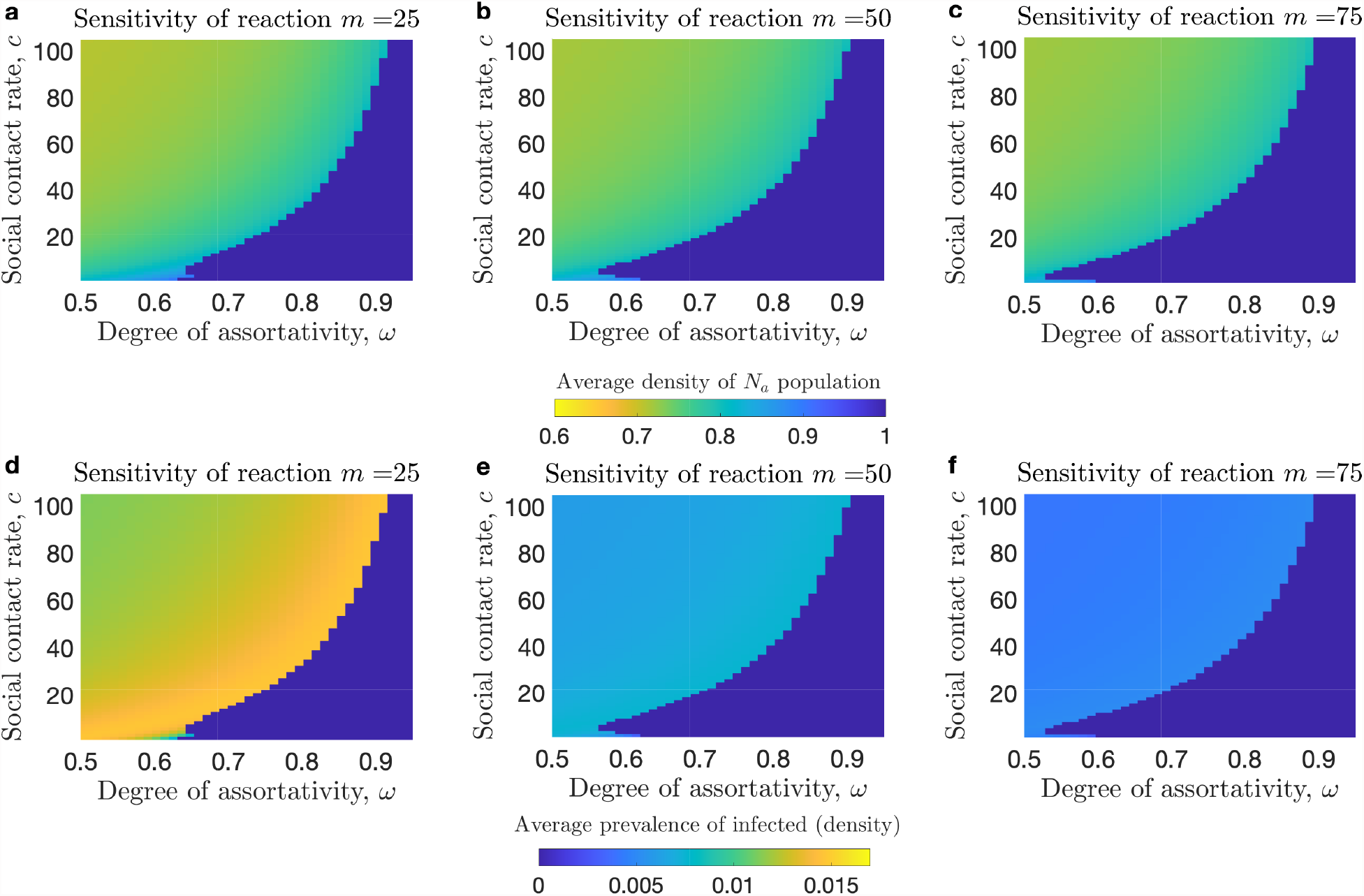
Impact of social contact rate and assortativity on the average endemic prevalence of infectious cases and average long-term opinion distribution. We consider the dynamics of the SIS system. **a, b**, and **c** show heat maps of the long term average density of the *N*_*a*_ population. **d, e**, and **f** show heat maps of the long term average infection prevalence. If the epidemic dynamics are periodic, then the average is taken over a period. **a** and **d** show scenarios with sensitivity of reaction to prevalence given by *m* = 25. **b** and **e** show scenarios with sensitivity of reaction to prevalence given by *m* = 50. **c** and **f** show scenarios with sensitivity of reaction to prevalence given by *m* = 75. The dark blue region in the top row and dark blue region in the bottom row denote the outcome where the population switched to opinion *a* and the disease becomes extinct. The infection rate of *N*_*b*_ individuals was set *β*_*b*_ = 1.5.

**Fig. 10.**
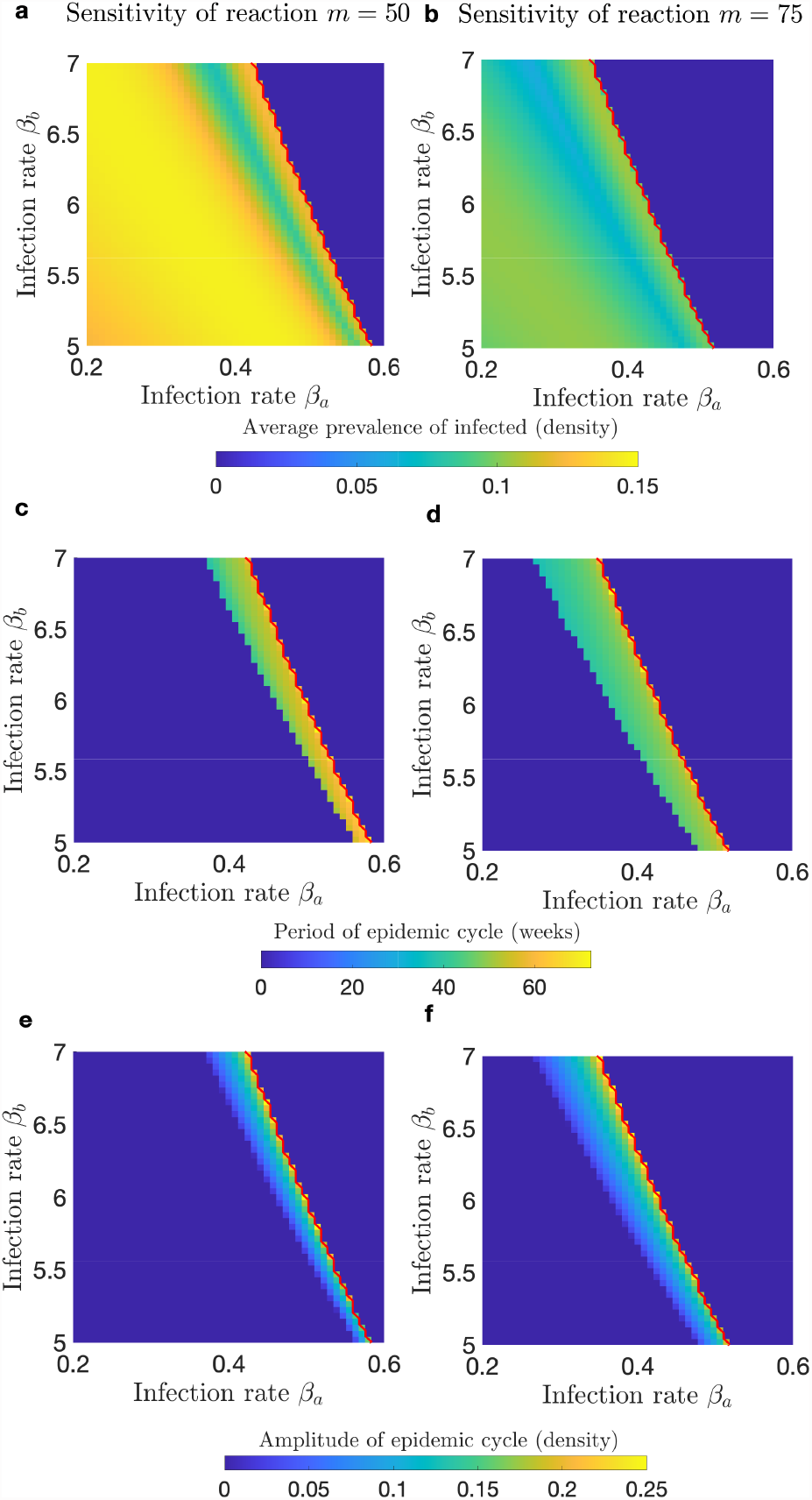
Impact of assortativity and sensitivity of reaction to the prevalence of infectious cases on the appearance of periodic epidemic dynamics. We consider the dynamics of the SIS system. We consider the dynamics of the SIS system. **a** and **b** show heat maps of the average prevalence. If the epidemic dynamics are periodic, then the average is taken over a period. **c** and **d** show heat maps of the period of the epidemic cycle. The period is equal to zero if the dynamics are stationary. **e** and **f** show heat maps of the amplitude of epidemic cycle. The amplitude is zero if the dynamics are stationary. **a, c**, and **e** show scenarios with sensitivity of reaction to prevalence given by *m* = 50. **b, d**, and **f** show scenarios with sensitivity of reaction to prevalence given by *m* = 75. The dark blue region above the red line denotes the outcome where the population switched to opinion *a* and the disease becomes extinct. The probability of switch to opinion *a* when no infectious cases are present is fixed *p*_*a*_ (0) = 0.28, the probability of switch to opinion *a* when the whole population is infected is fixed to *p*_*a*_ (1) = 0.6. Social contact rate is fixed *c* = 10.

Switching of the whole population to opinion *a* causes the disease to go extinct even when *R*_0_ *>* 1 for the opinion coexistence state. Our results indicate that higher sensitivity of the population to increasing prevalence, as reflected in high values of *m* and *p*_*a*_(1), will result in higher average densities of *n*_*a*_, and for some regions *n*_*a*_ = 1 (Figure 9 and Figure S6 in SI). The higher is the value of *m* the lower is the threshold value of *p*_*a*_(1) above which the population switches to opinion *a*. Moreover, if *p*_*a*_ is larger than a threshold value, the state *n*_*a*_ = 1 occurs for a wide range of sensitivity of the population to the prevalence, *m*. Should *p*_*a*_(1) exceed the threshold value significantly, the prevalence reduces considerably. Finally, high degree of assortativity in the population, on the one hand, leads to higher endemic prevalence. On the other hand, high assortativity leads to increase in the *p*_*a*_(1) − *m* subspace where the population switches to opinion *a*. We hypothesize that this is attributed to the positive effect assortativity has on infection transmission.

In addition to causing the population to switch to opinion *a* when a disease invades, the feedback between opinions competition and disease spread can induce sustained oscillatory epidemic dynamics (Figure 10). We investigated the conditions under which this may happen. We discovered that oscillatory dynamics mostly require a pronounced difference in epidemiological properties between individuals *N*_*a*_ and *N*_*b*_, such that when the whole population holds opinion *a*, the disease becomes extinct and if the whole population believes opinion *b* the disease persists. To show this, we plotted the amplitude of the epidemic cycle, its period and average value across an interval of infection rates values for two different sensitivities of the population reaction to the prevalence of infectious cases.

For a fixed value of infection rate of *N*_*b*_ individuals, *β*_*b*_ = 5.5, as infection rate of *N*_*a*_ individuals *β*_*a*_ increases initially, the endemic prevalence of infectious cases is constant in time, with the prevalence level increasing (Figure 11**a**). Once *β*_*a*_ increases past a threshold value, the constant endemic state is replaced by oscillatory dynamics, such that the average prevalence decreases as compared to the constant level it replaces (Figure 11**b**). As *β*_*a*_ increase further, the average prevalence, magnitude, and period of the cycle increase (Figure 11**c**). This pattern continues until the prevalence pushes the population to convert to opinion *a*, at which point the prevalence becomes zero and oscillatory dynamics disappear (Figure 10**d**).

**Fig. 11.**
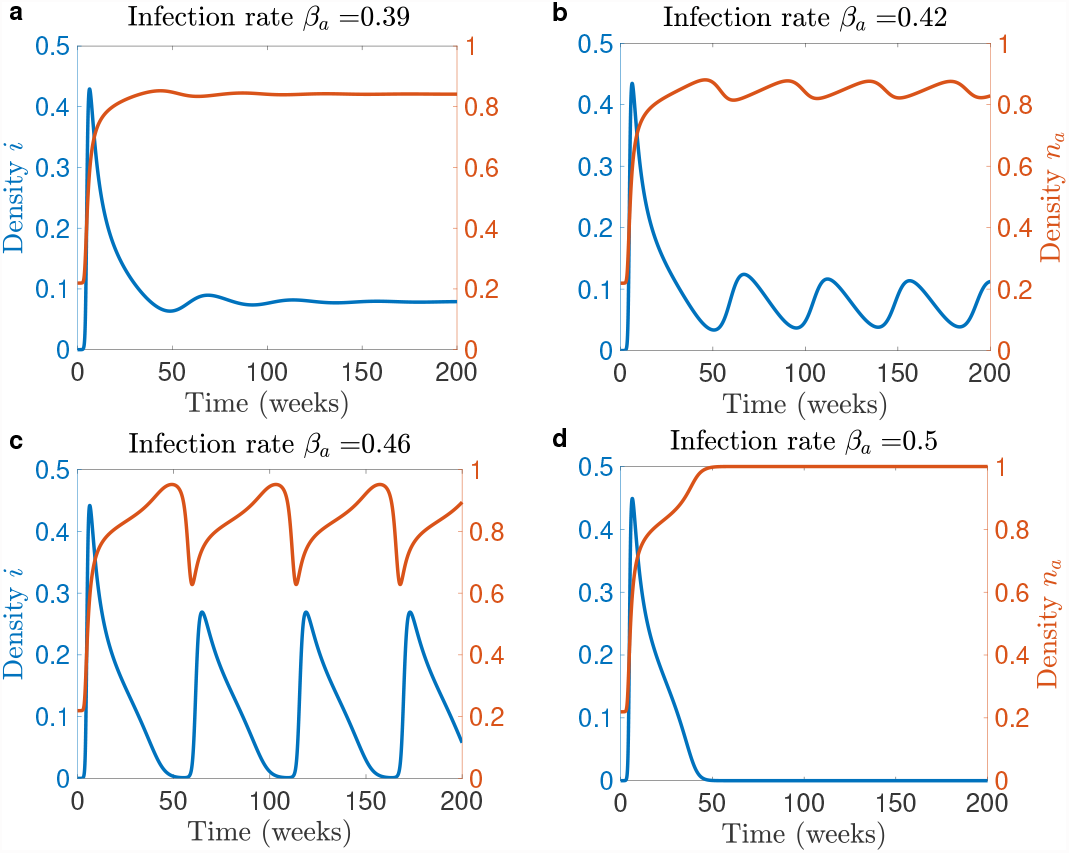
Sustained oscillatory dynamics resulting from feedback between disease dynamics and opinion dynamics. We consider the dynamics of the SIS system. Panels show time series for the prevalence of infectious cases and density of *N*_*a*_ population, *n*_*a*_ for different values of infection rate of *N*_*a*_ population, *β*_*a*_. The contact rate for information exchange is fixed *c* = 10, the probability of switch to opinion *a* when no infectious cases are present is fixed *p*_*a*_ (0) = 0.28, the probability of switch to opinion *a* per contact when the whole population is infected is fixed *p*_*a*_ (1) = 0.6, the constant that controls the growth of the switch rate to opinion *a* is fixed *m* = 75, and the infection rate of *N*_*b*_ individuals is fixed to *β*_*b*_ = 5.5.

To summarize, given a disease that follows SIS framework, adaptive behavior can lead to a number of qualitatively different outcomes. It can lead to the reduction of infection prevalence, appearance of sustained epidemic cycles, and complete eradication of the infection in conditions where the basic reproduction number would indicate that the infection will persist. Moreover, as the degree of assortativity increases and therefore, the basic reproductive number increases, the parametric region where opinion *a* becomes dominant becomes wider. Similar to the SIR model, the parameter region where oscillations arise is adjacent to the region where opinion *a* becomes the dominant opinion.

## Discussion

Using a model that couples opinion competition and infection spread, we investigated the effects of feedback between the two on epidemic dynamics. Our main findings were that the opinion distribution landscape can significantly influence the outcome of an epidemic. On the one hand, epidemic peaks can be reduced, and a population can be completely shifted into a health-positive state. On the other hand, damped or sustained oscillations of prevalence can appear as transmissibility of the infection increases. Parameters related to socializing dynamics such as social contact rate and degree of the assortative mixing by opinion were among the most important factors leading to the appearance of the above phenomena.

The influence of assortative mixing is two-fold. On the one hand, assortative mixing slows down the switching of opinions and therefore the possible reaction of the population to an epidemic. On the other hand, as the basic reproduction number increases as the assortative mixing increases, higher assortative mixing leads to higher incidence and therefore to a stronger reaction of the population, eventually even pushing the population into a state where the health-positive opinion is dominating. However, if assortativity is too high, its promoting effect on prevalence is not sufficient to help spread the health-protective opinion, and the population will experience a large epidemic peak. This effect on opinion spread is mitigated if the social contact rate is high.

Our model differs from earlier work incorporating awareness into epidemic modelling (2, 4) in that we consider both opinions as possibly attractive, such that a health-positive individuals may switch to a health-neutral opinion through contact with others who hold that opinion. This switching, which leads individuals to adopt a more risky health behavior, can therefore spread in the same way as health-positive behavior. In the papers (2, 4), awareness for the risks of infection decayed, when the infection was not present in the population, eventually leading to a completely unaware population. In contrast, in our model both opinions can co-exist in a steady state, also in a disease-free situation. The possibility of this outcome depends on the shape of opinion switch rate function. The potential of a stable co-existence of the two opinions implies that the impact of a new epidemic depends on the initial proportion of individuals with a health-positive opinion. Such an initial situation can be influenced, e.g., by educational interventions or other types of communication about future epidemic risks.

Appearance of oscillatory epidemic dynamics due to the feedback between health opinion dynamics and disease spread was observed both in the analysis of real world data (1, 5) and simulated trajectories produced by socio-epidemiological models (27, 28). In the present work, by means of considering changes in the dynamics across the parameter landscape, we gained insights into which properties of the system cause the appearance of oscillations. Pronounced difference between the carriers of two opinions in terms of infection rates as well as high average infection rate is one of the conditions for which oscillatory dynamics arise. Another important factor for the appearance of oscillations is a high rate of opinion exchange (as captured by the social contact rate) and high sensitivity of the population to prevalence. These two factors also contribute to the possibility of the population converting to the health positive opinion. In our experiments, the parametric regions where these two phenomena take place always appeared adjacent to each other.

The model can be extended to address present-day epidemic concerns, such as dynamics of infectious vaccine-preventable diseases. Vaccine uptake rate for well-known infectious diseases (e.g., measles, influenza) as well as for emerging ones (e.g., COVID-19) is fraught by reluctance of the part of the population to vaccinate (29–33). While circulation of vaccine uptake-endorsing opinions is subject to both communication from public health authorities as well as to interpersonal exchanges (30, 34, 35), the circulation of anti-vaccination sentiment depends on social norms within the local network and interpersonal communications within the network (34–36). The models that considered the role of interpersonal communications on the vaccination uptake and its effect on epidemic dynamics (27, 28), while coupling vaccination strategies with the population epidemic state, modeled the growth of the vaccinating population contingent on the presence of the disease, while its opposite, non-vaccinating sentiment, did not depend on the population state. Our framework which allows for symmetric treatment of health-positive and health-neutral sentiments is well-suited for investigation of vaccination opinion dynamics with or without the disease.

Our framework can bring interesting qualitative insights for the dynamics of a vaccine preventable disease characterized by waning immunity (e.g. measles, pertussis, influenza). In the conditions of waning immunity, it is highly important to keep up consistently high vaccination uptake rate if not to eradicate the disease, at least to avoid the overcrowding of the health care system. Another important consideration, in the context of infectious diseases characterized by waning immunity, is the process of waning and boosting of immunity which can cause pronounced oscillation dynamics (37). Therefore, for infectious diseases characterized by waning and boosting of immunity, presence of adaptive behavior with respect to vaccination, can give rise to rich dynamics highly relevant for the efforts of health authorities.

In this work, we assumed that the social exchange does not necessarily require physical contacts (interactions that have a probability of infection transmission), i.e.… in a situation where the physical contact may decrease, the information exchange and thus, opinion dynamic will proceed unimpeded. However, in real life, at least some of the social contacts will terminate if the physical contact rate is reduced. Thus, if health-positive individuals practice social distancing then opinion dynamics and subsequently epidemic dynamics will be altered in a number of ways that may not necessarily benefit the population. For example, given a reduction of social contact rate for the health-positive individuals, it may be necessary they are present at a higher proportion, in order to maintain steady presence in the population.

Our simple model has rich dynamics, appearance of which depends on the functional responses and parameter values. For example, as our analyses have shown, the shape of the functional response plays a key role in the dynamics of health opinions/behaviors and subsequently in epidemic dynamics. Therefore, to be able to use the model for qualitative and quantitative predictions it is paramount to accurately identify functional representations for the opinion switch rates and for behavioral response to the epidemic spread. Having these at hand will enable the design of information interventions to be well-tailored to the specific time frame of the epidemic.

## Materials and Methods

The system of ordinary equations (6) describes the coupled dynamics of infection spread and opinion competition.

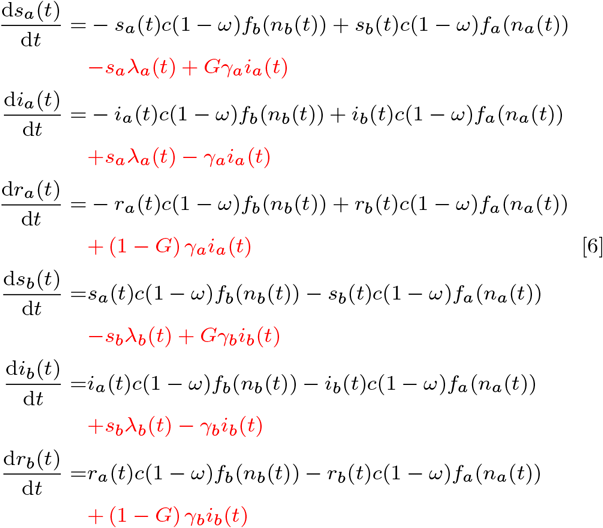

where

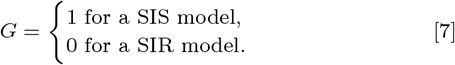

and *λ*_*a*_ and *λ*_*b*_ are specified by equations (2).

### Model code

The model was implemented in MATLAB R2021b (38). The code producing the analyses and figures for this study is available at https://github.com/aiteslya/TwoOpinion (39).

## Data Availability

All data produced are available online at the repository on Github.

https://github.com/aiteslya/TwoOpinion

## ACKNOWLEDGMENTS

Please include your acknowledgments here, set in a single paragraph. Please do not include any acknowledgments in the Supporting Information, or anywhere else in the manuscript.

