## Supplementary materials for "The effect of competition between health opinions on epidemic dynamics"

##### This PDF file includes:

Supplementary text
Figs. S1 to S6

### Supporting Information Text

#### Equilibria and their stability of opinion competition system

To find the equilibria in an opinion competition model without epidemic dynamics, we set $i_a = r_a = i_b = r_b = 0$. Since the population density is constant and adds up 1, we replace $s_b = 1 - s_a$, set the equation describing the dynamics of $s_a$ to zero, cancel term $c(1-\omega)$ and solve for $s_a$. We obtain the following equation, solutions of which are equilibria of system (6):

$$- s_a f_b(s_b(t)) + s_b f_a(s_a(t)) = 0 \ . \qquad [1]$$

Since $f_a(0) = f_b(0) = 0$, there always exist two equilibria: $(s_a^*, 1 - s_a^*) = (1, 0)$ and $(s_a^*, 1 - s_a^*) = (0, 1)$. Their local and global stability depend on the shape of the opinion switch rate functions.

**Linear switch rate function.** If the switch rates are linear (i.e. given by Eq. (1) with $k = 1$, $\theta_a = 0$, and $\theta_b = 0$, Eq. (1) reduces to

$$- p_b s_a s_b + p_a s_a s_b = 0 \ . \qquad [2]$$

If $p_a \neq p_b$, the only possible solutions are $(s_a, s_b) = (1, 0)$ and $(s_a, s_b) = (0, 1)$ and no permanent co-existence of opinions can take place. The co-existence of opinions is possible only when $p_a = p_b$, and then any combination of $s_a$ and $s_b$ such that $s_a + s_b = 1$ will be a steady state. While mathematically correct, this solution is not biologically viable.

The linearization of the system about an equilibrium $(s_a^*, 1 - s_a^*)$ yields:

$$J_1 = (1 - 2s_a^*)(p_a - p_b) \ . \qquad [3]$$

If $p_a > p_b$, then (1,0) is globally asymptotically stable and (0,1) is unstable. If $p_a < p_b$, then (1,0) is unstable and (0,1) is globally asymptotically stable. If $p_a = p_b$, all equilibria are stable.

**Saturating switch rate function.** If the switch rate functions are saturating (i.e. given by Eq. (1) with $k = 1$, $\theta_a > 0$, and $\theta_b > 0$, Eq. (1) reduces to

$$- \frac{p_b}{1 + \theta_b(1 - s_a)} + \frac{p_a}{1 + \theta_a s_a} = 0 \ . \qquad [4]$$

Solve for $s_a$ to obtain

$$\bar{s}_a = \frac{p_a - p_b + p_a\theta_b}{p_b\theta_a + p_a\theta_b} \ . \qquad [5]$$

In order for $0 < \bar{s}_a < 1$ to hold, the following condition ("co-existence of opinions" condition) should hold:

$$\frac{p_a}{p_b} > \frac{1}{1 + \theta_b} \ , \qquad [6a]$$

$$\frac{p_a}{p_b} < 1 + \theta_a \ . \qquad [6b]$$

The intuitive interpretation of the above condition is that in order for the permanent co-existence of opinions to occur, the size of the probabilities of switch per contact to either of the opinions should be relatively close to each other.

The linearization of system (6) about an equilibrium $(s_a^*, 1 - s_a^*)$ yields:

$$J_2 = (1 - s_a)D(s_a) - s_a D(s_a) + s_a(1 - s_a)\frac{\partial D(s_a)}{\partial s_a} \ , \qquad [7]$$

where

$$D(s_a) = \frac{-p_b}{1 + \theta_b(1 - s_a)} + \frac{p_a}{1 + \theta_a s_a} \ . \qquad [8]$$

At $(s_a^*, 1 - s_a^*) = (1, 0)$, Eq. (11) reduces to

$$J_2\big|_{s_a^*=1} = -D(1) \ . \qquad [9]$$

Therefore, $(s_a^*, 1 - s_a^*) = (1, 0)$ is unstable when inequality (6a) holds and stable otherwise.

At $(s_a^*, 1 - s_a^*) = (0, 1)$, Eq. (11) reduces to

$$J_2\big|_{s_a^*=1} = D(0) \ . \qquad [10]$$

Therefore, $(s_a^*, 1 - s_a^*) = (0, 1)$ is unstable when inequality (6b) holds and stable otherwise.

Thus, both $(1, 0)$ and $(0, 1)$ are unstable when the co-existence of opinions equilibria exists.

Observe that at $D(\bar{s}_a) = 0$. Therefore,

$$J_2\big|_{s_a=\bar{s}_a} = \bar{s}_a(1 - \bar{s}_a)\frac{\partial D(s_a)}{\partial s_a}\bigg|_{s_a=\bar{s}_a} \ . \qquad [11]$$

When the co-existence opinion equilibrium exists, inequality (6a) holds. Therefore, $\frac{\partial D(s_a)}{\partial s_a} < 0$, and thus, $J_2\big|_{\bar{s}_a} < 0$. It follows that, the co-existence equilibrium $(\bar{s}, 1 - \bar{s})$, where $\bar{s}$ is given by Eq. 5, is locally asymptotically stable.

**Sigmoidal switch rate function.** If the switch rate function is sigmoidal (i.e. given by Eq. (1) with $k > 1$, $\theta_a > 0$, and $\theta_b > 0$), Eq. (1) becomes:

$$-s_a \frac{p_b(1-s_a)^k}{1+\theta_b(1-s_a)^k} + (1-s_a)\frac{p_a s_a^k}{1+\theta_a s_a^k} = 0 \; . \tag{12}$$

At the co-existence of opinions equilibrium $(\bar{s}_a, 1-\bar{s}_a)$, such that $0 < \bar{s}_a < 1$, the following condition must hold:

$$\mathcal{F}(s_a) = -\frac{p_b(1-s_a)^{k-1}}{1+\theta_b(1-s_a)^k} + \frac{p_a s_a^{k-1}}{1+\theta_a s_a^k} = 0 \; . \tag{13}$$

Set

$$\Phi_1(s_a) = \frac{p_b(1-s_a)^{k-1}}{1+\theta_b(1-s_a)^k} \; , \tag{14a}$$

$$\Phi_2(s_a) = \frac{p_a s_a^{k-1}}{1+\theta_a s_a^k} \; . \tag{14b}$$

Note that $\mathcal{F} = -\Phi_1 + \Phi_2$, $\Phi_1(s_a > 0$, and $\Phi_2(s_a) > 0$ for $0 < s_a < 1$. Since $\Phi_1(0) = \frac{p_b}{1+\theta_b} > 0$, $\Phi_1(1) = 0$, $\Phi_2(0) = 0$, and $\Phi_2(1) = \frac{p_a}{1+\theta_a} > 0$, then Eq. (13) has at least one solution $(\bar{s}_a, 1-\bar{s}_a)$, such that $0 < \bar{s}_a < 1$.

Let us consider next the stability of the equilibria. The linearization of the system around an equilibrium $(s_a^*, 1-s_a^*)$ yields:

$$J_3 = (1-s_a^*)\mathcal{F}(s_a^*) - s_a^*\mathcal{F}(s_a^*) + s_a^*(1-s_a^*)\left(-\frac{\partial\Phi_1(s_a^*)}{\partial s_a^*} + \frac{\partial\Phi_2(s_a^*)}{\partial s_a^*}\right) \; . \tag{15}$$

At $(s_a^*, 1-s_a^*) = (1, 0)$, Eq. (15) reduces to

$$J_3|_{s_a^*=1} = -\mathcal{F}(1) < 0 \; . \tag{16}$$

Thus, the equilibrium $(s_a^*, 1-s_a^*) = (1, 0)$ is always locally asymptotically stable.

At $(s_a^*, 1-s_a^*) = (0, 1)$, Eq. (15) reduces to

$$J_3|_{s_a^*=0} = \mathcal{F}(0) < 0 \; . \tag{17}$$

Thus, the equilibrium $(s_a^*, 1-s_a^*) = (0, 1)$ is always locally asymptotically stable.

At $(s_a^*, 1-s_a^*) = (\bar{s}_a, 1-\bar{s}_a)$, such that $0 < s_a < 1$, Eq. (15) reduces to

$$J_3|_{s_a^*=\bar{s}_a} = s_a^*(1-s_a^*)\left(-\frac{\partial\Phi_1(s_a^*)}{\partial s_a^*} + \frac{\partial\Phi_2(s_a^*)}{\partial s_a^*}\right) \; . \tag{18}$$

The sign of the Jacobian is determined by signs of the derivatives of $\Phi_1$ and $\Phi_2$, and these themselves depend on the magnitudes of $p_a$, $p_b$, $k$, $\theta_a$, and $\theta_b$.

If there is a single co-existence of opinions equilibrium, then by general theory of the dynamical systems, it is unstable. If there is more than one co-existence of opinions equilibrium, then there is an odd number of them, $2g+1$, $g \in \mathbb{Z}$ such that $g+1$ of these equilibria are unstable and $g$ equilibria are locally stable.

#### Basic reproduction number

In Figure S1 we investigated how the basic reproduction number $R_0$ changes with changing social contact rate $c$ and assortativity degree $\omega$ for three settings of the ratio $p_a/p_b$: 0.8, 1, and 1.25, when the infection rate of $N_b$ individuals is set $\beta_b = 1.5$, the value used to investigate the dynamics of the SIS model.

We also investigate variation of the basic reproduction number in the parameter space $\beta_a - \beta_b$ that was considered in the main analysis (Figure S2). We give the results for the parameter region used for the SIR model and for the SIS model.

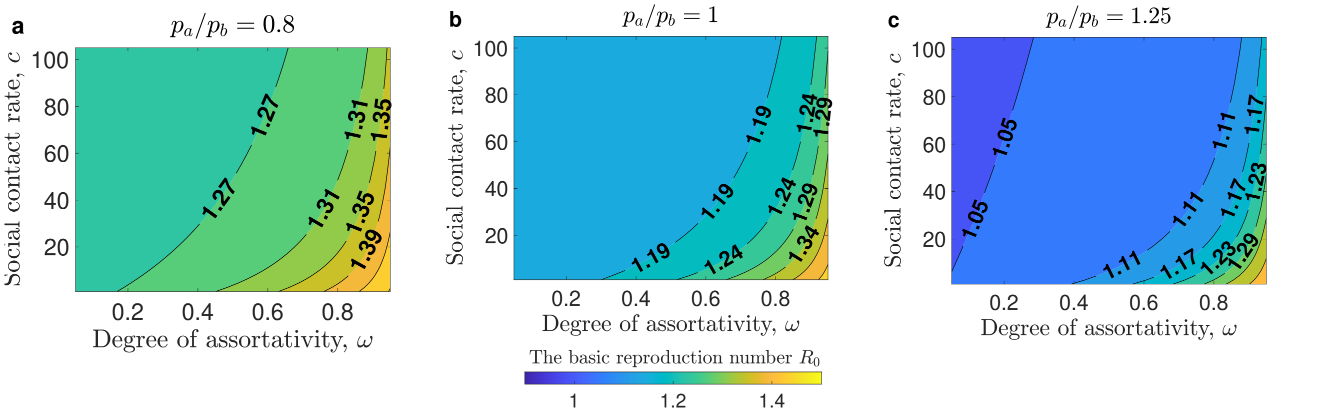


**Fig. S1. Impact of mixing patterns on basic reproduction number $R_0$.** **a**, **b**, and **c** show contour maps of $R_0$ as a function of the social contact rate $c$ and the assortativity $\omega$. **a** For $p_a/p_b = 0.8$ the initial distribution of opinions is $(s_a(0), s_b(0)) = (0.35, 0.65)$. **b** For $p_a/p_b = 1$ we have $(s_a(0), s_b(0)) = (0.5, 0.5)$. **c** For $p_a/p_b = 1.25$ we have $(s_a(0), s_b(0)) = (0.65, 0.35)$. The infection rate of susceptible individuals holding opinion $a$ is fixed $\beta_b = 1.5$, the value used to investigate the dynamics for the SIS system.

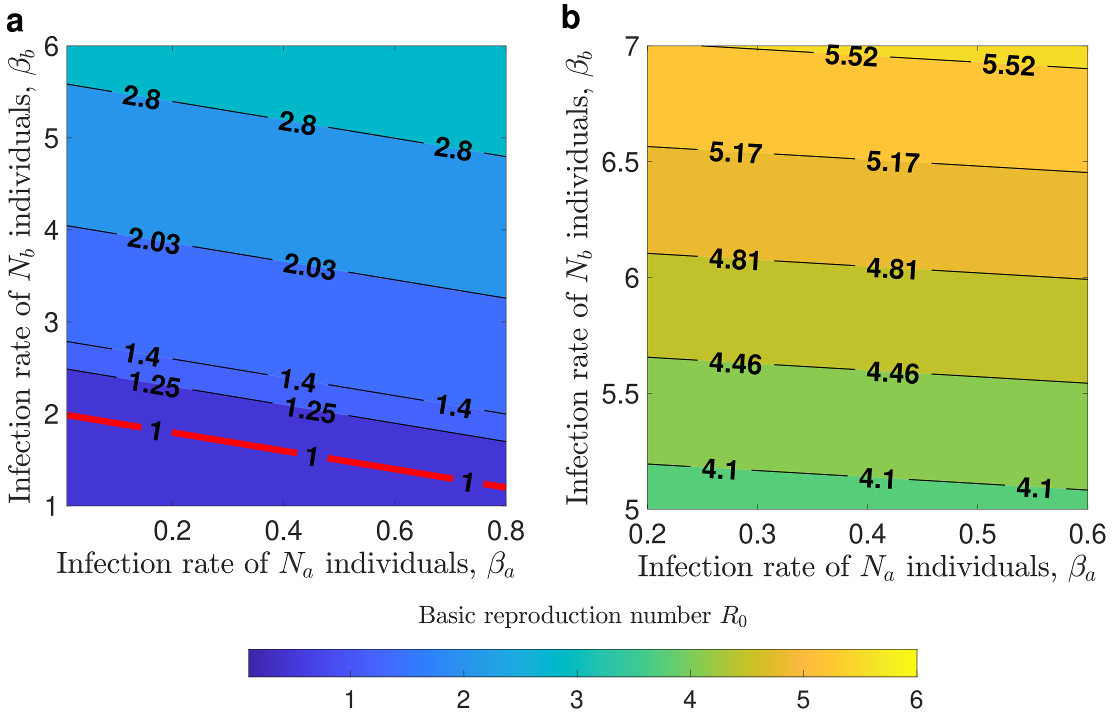


**Fig. S2. Basic reproduction number $R_0$ in $\beta_a - \beta_b$ space.** Contour plots show the basic reproductive number for different regions in $\beta_a - \beta_b$ space. **a** shows the region used for SIR simulations, **b** shows the region used for SIS simulations. The assortativity was set $\omega = 0$, the social contact rate was set $c = 40$.

#### Multiple epidemic peaks under SIR framework

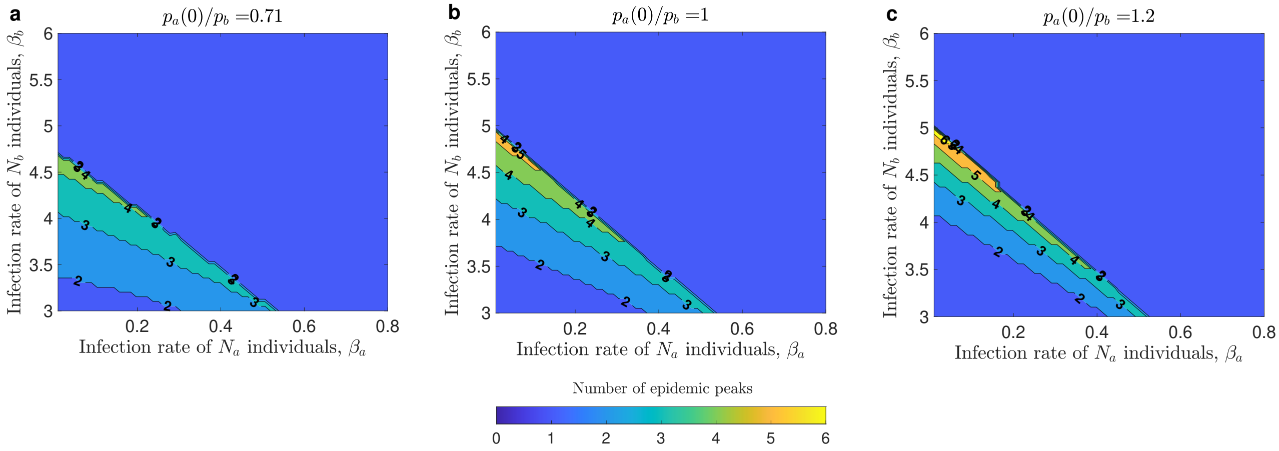


**Fig. S3. Appearance of multiple epidemic peaks as a result of the feedback between the disease dynamics and opinion dynamics**. We consider the dynamics of the SIR system. Contour plots show the number of the prevalence peaks for different values of infection rates of population $N_a$, $\beta_a$ and of population $N_b$, $\beta_b$, given three different ratios of probabilities of switch $p_a/p_b$ at the start of the pandemic. **a** shows the contour plot for $p_a/p_b = 0.71$, **b** for $p_a/p_b = 1$, and **c** for $p_a/p_b = 1.2$. The contact rate for information exchange is fixed $c = 40$ and the sensitivity of reaction to changes in the prevalence of infectious cases is set to $m = 75$.

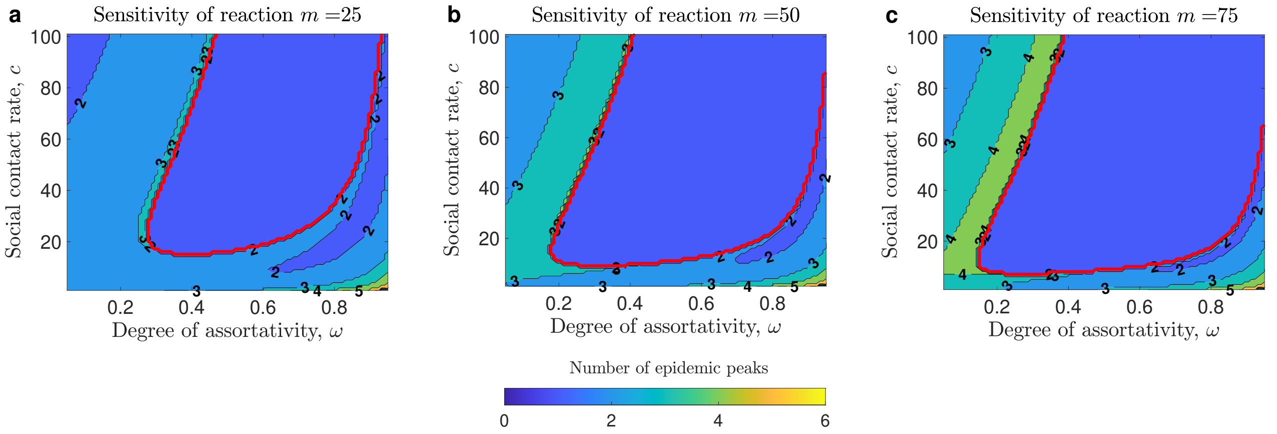


**Fig. S4. Appearance of multiple epidemic peaks as a result of the feedback between the disease dynamics and opinion dynamics**. We consider the dynamics of the SIR system. Contour plots show the number of the prevalence peaks in $\omega - c$ space for different sensitivities of population reaction to prevalence. **a** shows the contour plot for $m = 25$, **b** for $m = 50$, and **c** for $m = 75$. The infection rate for $N_a$ individuals is fixed $\beta_a = 0.1$ and the infection rate for $N_b$ individuals is fixed $\beta_b = 4$.

#### Peak prevalence and peak density of population $N_a$ under SIR

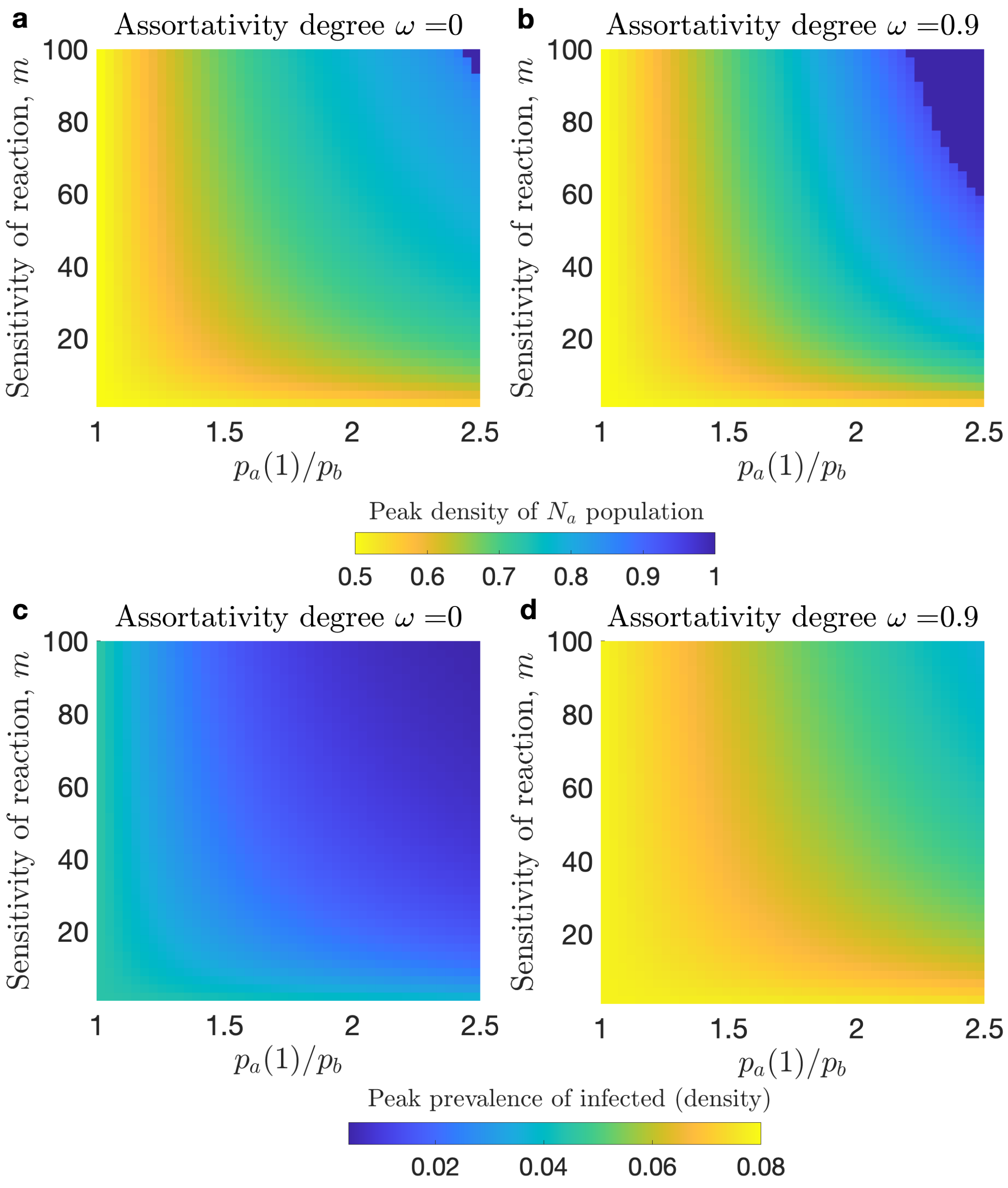


**Fig. S5. Impact of assortativity and sensitivity of reaction to the prevalence on the peak prevalence of infectious cases and long-term opinion distribution**. We consider the dynamics of the SIR system. **a** and **b** show heat maps of the peak density of $N_a$ population. **c** and **d** show heat maps of the peak prevalence. **a** and **c** show scenarios with no assortative mixing in the population. **b** and **d** show scenarios with high assortativity by opinion. Bright yellow region on **a** and **c** denote the outcome where the population switched to opinion $a$. As assortativity increases this region expands.

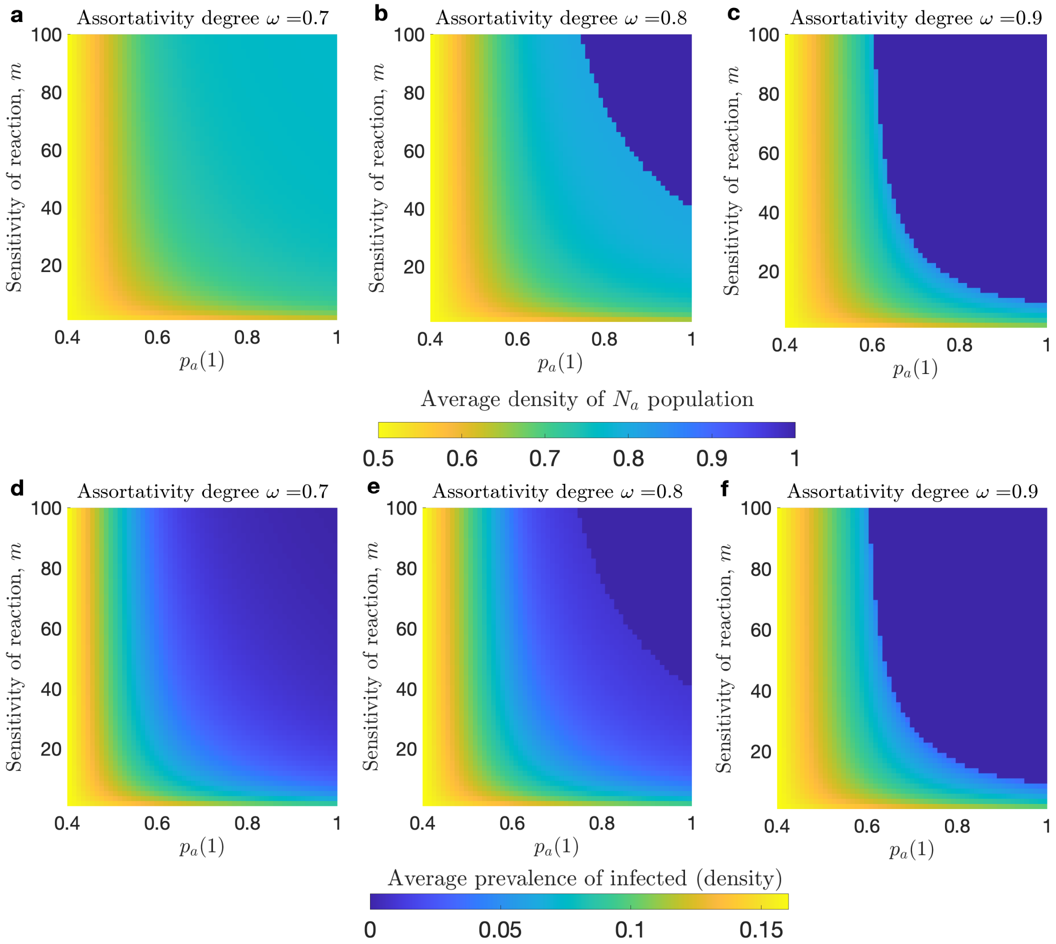


**Fig. S6. Impact of assortativity and sensitivity of reaction to the prevalence of infection on the endemic prevalence of infectious cases and long-term opinion distribution**. We consider the dynamics of the SIS system. **a**, **b**, and **c** show heat maps of the average density of $N_a$ population. **d**, **e**, and **f** show heat maps of the average prevalence. **a** and **d** show scenarios with assortative degree fixed $\omega = 0.7$. **b** and **e** show scenarios with assortative degree fixed $\omega = 0.8$. **c** and **f** show scenarios with assortative degree fixed $\omega = 0.9$. The blue region on **a**-**c** denotes the outcome where the population switched to opinion $a$. As assortativity increases this region expands.

81 # References
